# Impact of Early Treatment on Symptom Improvement and Procedural Events among Men with BPH and Bothersome Lower Urinary Tract Symptoms: A Contemporary Analysis of the American Urological Association Quality (AQUA) Registry

**DOI:** 10.64898/2026.06.08.26355194

**Authors:** John Ernandez, Allison Najafi, Claus G. Roehrborn, Lori B. Lerner

## Abstract

**PURPOSE:** As the armamentarium of BPH therapies continues to expand, it remains imperative to maximize patient satisfaction and minimize decisional regret. We sought to determine the impact of time from BPH diagnosis to index treatment on symptom improvement and subsequent procedural events.

**MATERIALS AND METHODS:** We queried the American Urological Association Quality Registry for men ≥ 40 years old with BPH, available IPSS data, and no receipt of prior BPH treatment. Index treatment included medication, surgery, or minimally invasive surgical therapy (MIST). Outcomes included IPSS over 3 years of follow-up, change in percentage of mild lower urinary tract symptoms (LUTS) by 3 months, and time to procedural event. Patients were stratified by time from index diagnosis to treatment by <12 months, 1-3 years, and >3 years. Outcomes were compared across time-to-treatment cohorts with appropriate statistical tests with *p* < 0.05 as significant.

**RESULTS:** 43,919 patients met criteria with 19,642 pursuing treatments. Patients pursued treatment at comparably lower baseline IPSS compared to prior prospective series. Patients undergoing surgery and MIST had significantly higher baseline IPSS, while medical comorbidities were significantly more common among men initiating pharmacotherapy. Early surgery and MIST were associated with significant improvement in IPSS within 6-12 months and an increase in mild LUTS by 3 months. All forms of early treatment were associated with delayed time to procedural events, including catheterization and fulguration.

**CONCLUSION:** Early procedural intervention for BPH is associated with early symptom improvement and delayed time to procedural events among real-world, contemporary practice.

## INTRODUCTION

Bladder outlet obstruction due to benign prostatic hyperplasia (BPH) can result in clinically significant lower urinary tract symptoms (LUTS) in 50% of men with BPH.^1^ Additionally, 75 – 90% of men will have histologic evidence of BPH by age 80.^2^ Accordingly, the prevalence of bothersome LUTS is expected to increase with the aging U.S. population.^1^

The armamentarium of available therapies for BPH continues to expand. For patients bothered by their LUTS, medical treatment including use of α-blockers, 5α-reductase inhibitors (5ARIs), or combination therapy, remains the mainstay of first-line treatment. While pharmacotherapy can improve urinary flow, reduce risk of urinary retention, and delay progression to surgery,^3–4^ patients with either bothersome side effects^5–6^ or absolute indications for surgical intervention,^7^ may pursue surgical treatment. Advances in technology have resulted in an expansion of available surgical therapies for BPH, including laser enucleation, photoselective vaporization (PVP), and Aquablation, which have been demonstrated to be safe, effective, and durable treatments.^8–10^ Additionally, minimally invasive surgical therapies (MISTs) have become increasingly available and offer patients more favorable side effect profiles.^11–12^

While quality of life generally worsens as BPH progresses,^13^ unmet expectations after treatment have also been shown to be associated with decisional regret.^14–15^ Additionally, changes in patient-reported International Prostate Symptom Score (IPSS) have been associated with treatment satisfaction after BPH treatment.^16^ Accordingly, management of LUTS relies on appropriately timing elective treatment to maximize patient satisfaction and minimize regret. The American Urological Association Quality (AQUA) Registry, which collects contemporary clinical data from participating practices across the U.S., allows for a unique opportunity to determine the impact of treatment timing on patient-reported symptoms.

We sought to determine if time from BPH diagnosis to index medical or surgical treatment impacted symptomatic improvement and subsequent procedural events within the AQUA Registry. We hypothesized that patients who delayed treatment would be more likely to report poorer symptom improvement and shorter time to procedural events due to both disease progression and symptomatic bother.

## MATERIALS AND METHODS

### Cohort

The AQUA Registry was queried to identify patients ≥ 40 years old with relevant diagnosis code(s) for BPH on ≥ 2 clinical encounters between July 1, 2015, and July 31, 2023. Patients had to have available IPSS scores at or prior to index treatment and at least 1 IPSS score available after treatment, two valid clinic notes, at least one year of follow-up, no prior urologic surgery, and no receipt of BPH or prostate cancer treatment. Should patients have multiple IPSS scores recorded per date, the highest score was recorded for analysis. If more than one IPSS score was recorded between follow-up end points, the score closest to the end point of interest was recorded.

### Covariates

Baseline clinicodemographic covariates included age, race, ethnicity, region of care received based on U.S. Census, IPSS score, prostate specific antigen (PSA), and comorbidities. Treatments were categorized by watchful waiting, medical treatment, MIST, and surgical treatment. Medical treatment included use of alpha-blockers, 5ARIs, phosphodiesterase-5 inhibitors, and any combination of drug categories. Patients using alpha blockers for hypertension, 5ARIs for male-pattern baldness, or PDE5Is for erectile dysfunction were excluded. Overactive bladder (OAB) medications included anticholinergics and beta-3 agonists. MISTs included Urolift and Rezum. Surgical treatments included transurethral resection of prostate (TURP), PVP, laser enucleation, and Aquablation.

### Outcomes

IPSS scores were recorded for up to 3 years after index treatment at time points of 3, 6, 12, 24, and 36 months. Patients were stratified based on time from diagnosis to index treatment (referred to as “time to treatment” or TTT), with TTT categories including <12 months, 1-3 years, and >3 years. Early symptom improvement was defined as an increase in proportion of mild LUTS by IPSS score by 3 months after index treatment. Catheterization and bladder irrigation after index treatment were selected as indicators of disease progression and felt to contribute to burden of care. They were termed “procedural events”. Fulguration and clot evacuation were also recorded as procedural events for patients pursuing MIST or surgery. Time to procedural events were recorded for each treatment type. Time to addition of OAB medications or re-treatment with surgery or MIST were also recorded for patients pursuing index medical treatment. Time to surgical re-treatment for patients pursuing index MIST or surgery was also recorded.

### Statistical Analysis

Baseline covariates were summarized using mean and standard deviation, median and interquartile range (IQR), or frequency counts with percentages and compared across treatment groups via Pearson chi-squared test for categorical variables and Kruskal-Wallis test for continuous variables. Overall comparisons across TTT categories for change in IPSS over follow-up were tested via Kruskal-Wallis tests, while pairwise comparisons between and within TTT categories were tested via Mann-Whitney U test with appropriate Bonferroni corrections. Comparisons of early symptom improvement at 3 months and 30-day and 12-month event frequencies across TTT categories were made via Pearson chi-squared tests. Overall comparisons of mean time to retreatment across TTT categories were made via one-way Welch ANOVA with pairwise comparisons between TTT categories made with two-way Welch *t-*test with Bonferroni correction. Statistical analyses were performed using Graphpad Prism 10.6.1 (GraphPad Software). Figures 1 and 2 were generated in assistance with Claude Sonnet 4.6 (Anthropic) and reviewed by authors to ensure accurate representation of data. All tests were two-sided with *p*-values < 0.05 considered statistically significant.

**Figure 1.**
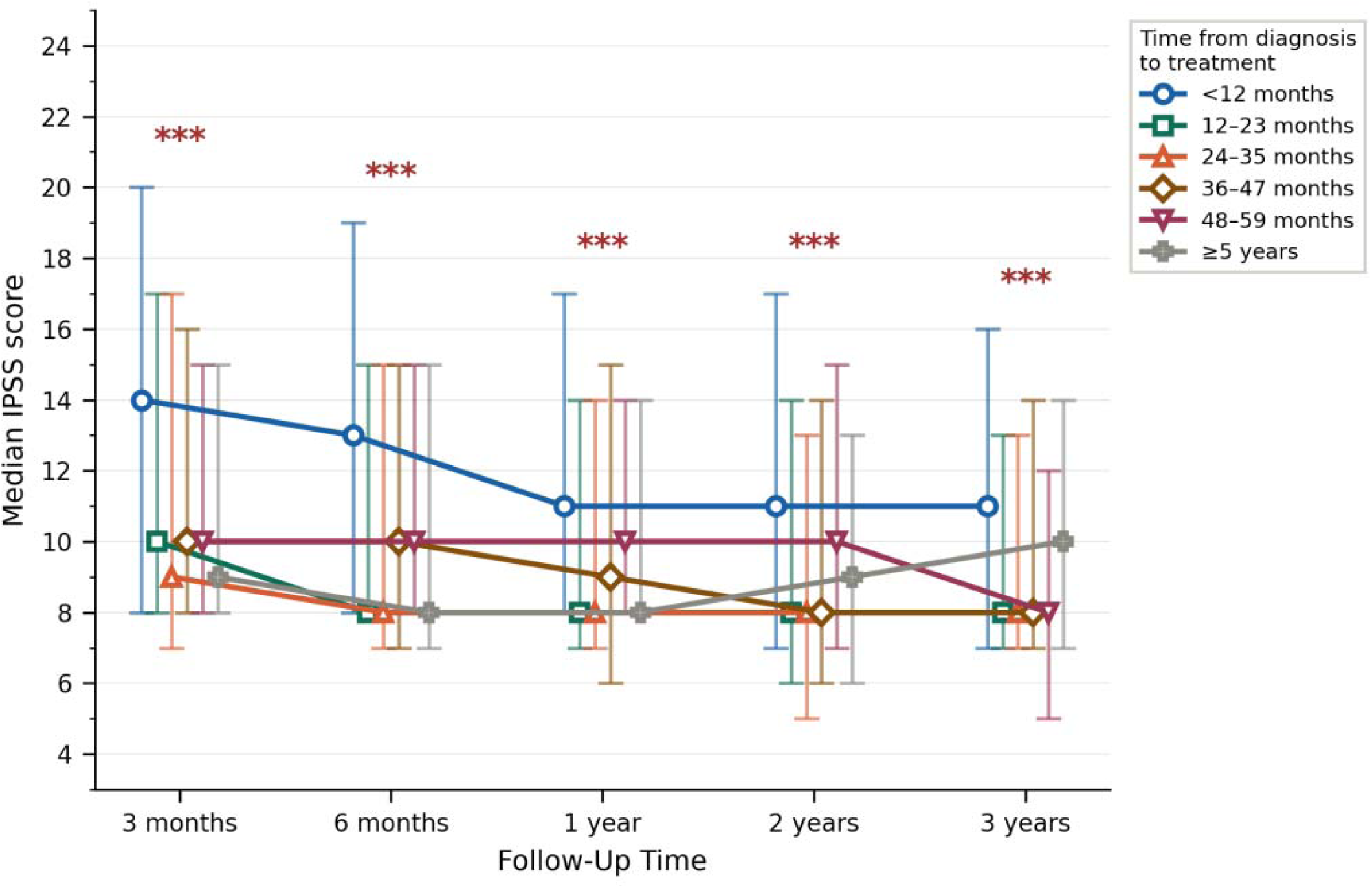
Median IPSS score over follow-up stratified by time from diagnosis to medical treatment of BPH. *** denotes significant between-group differences at *p* < 0.001.

**Figure 1b.**
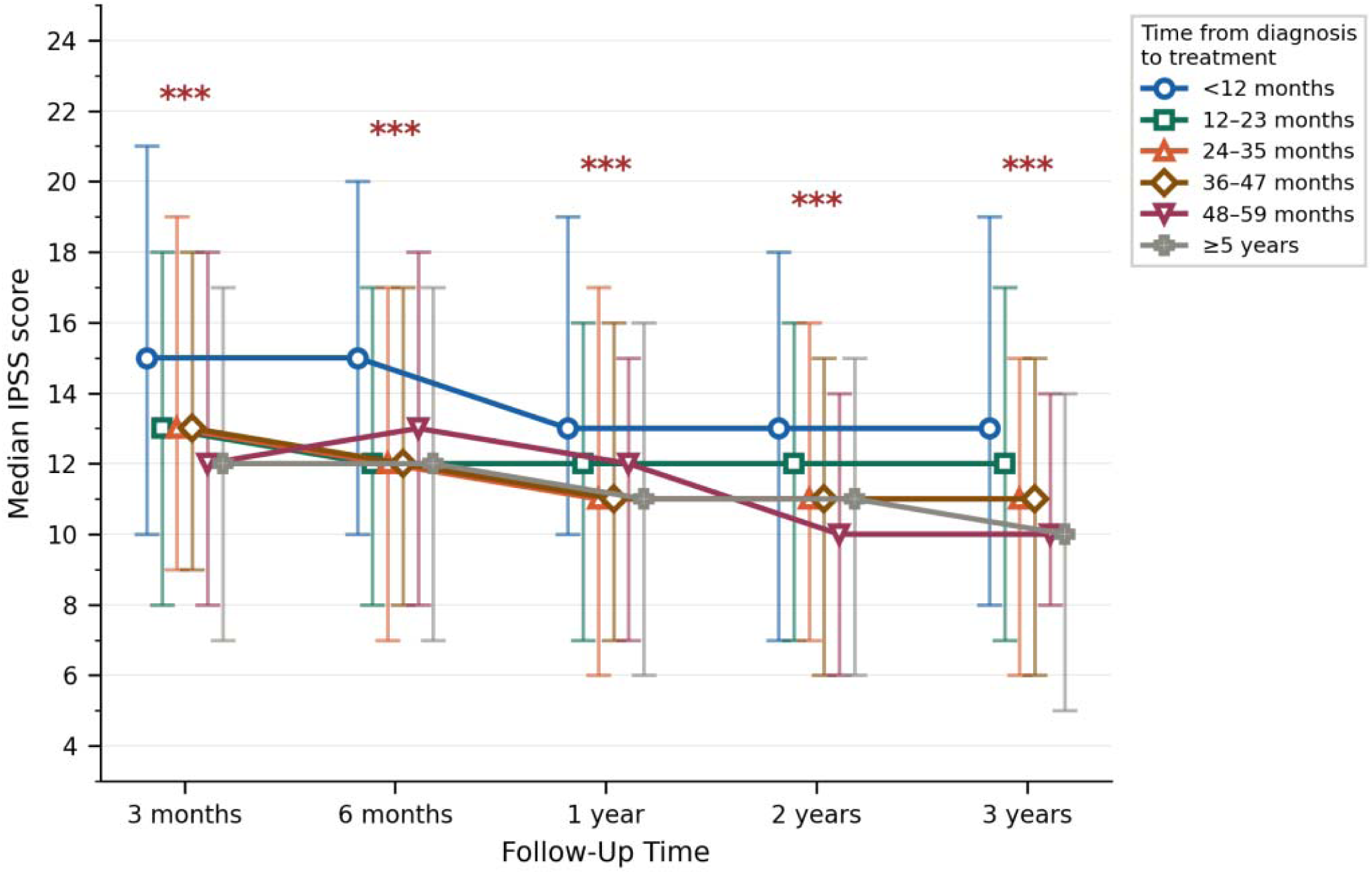
Median IPSS score over follow-up stratified by time from diagnosis to MIST for BPH. *** denotes significant between-group differences at *p* < 0.001.

**Figure 1c.**
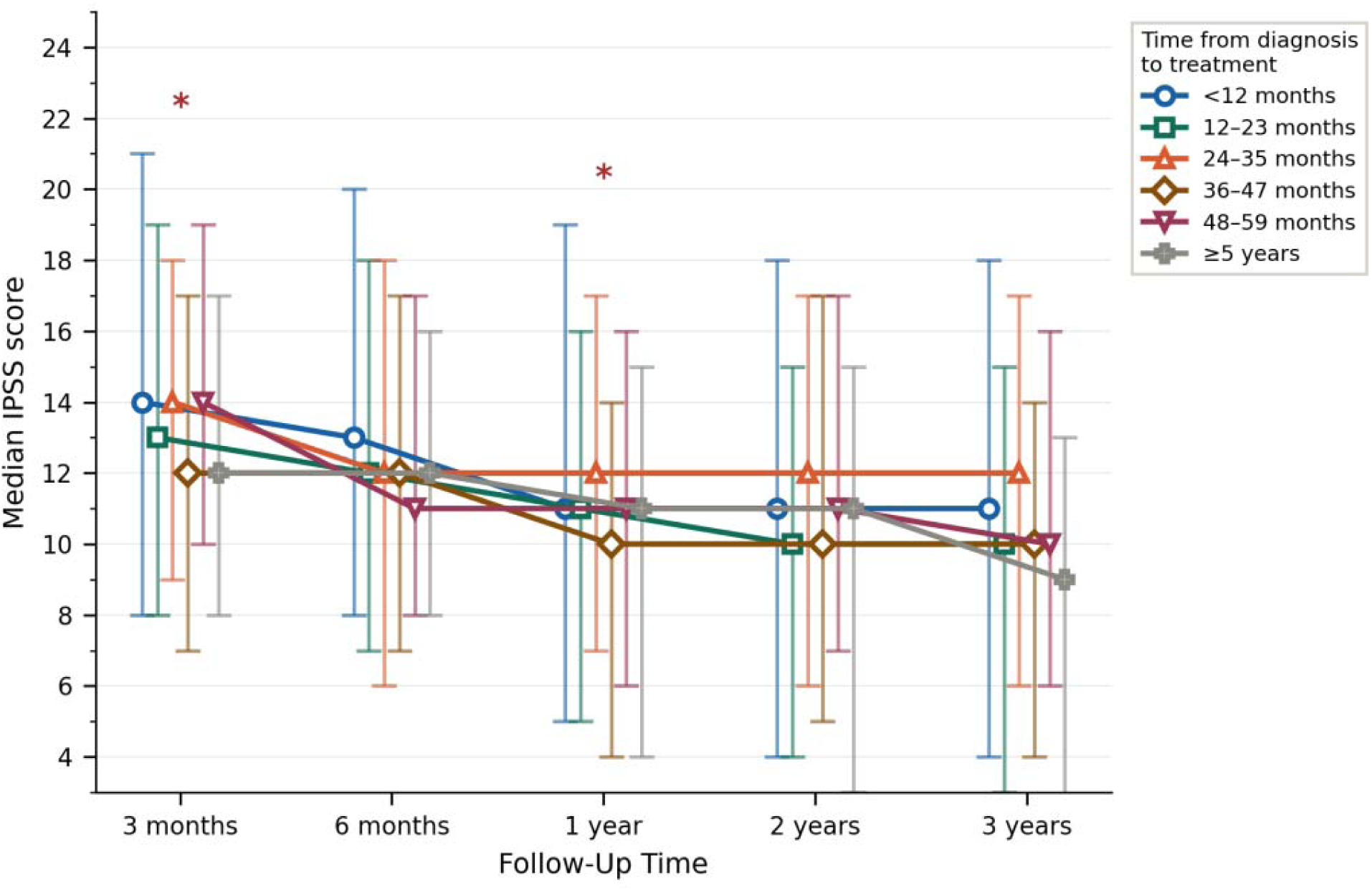
Median IPSS score over follow-up stratified by time from diagnosis to surgical treatment of BPH. * denotes significant between-group differences at *p* < 0.05.

**Figure 2.**
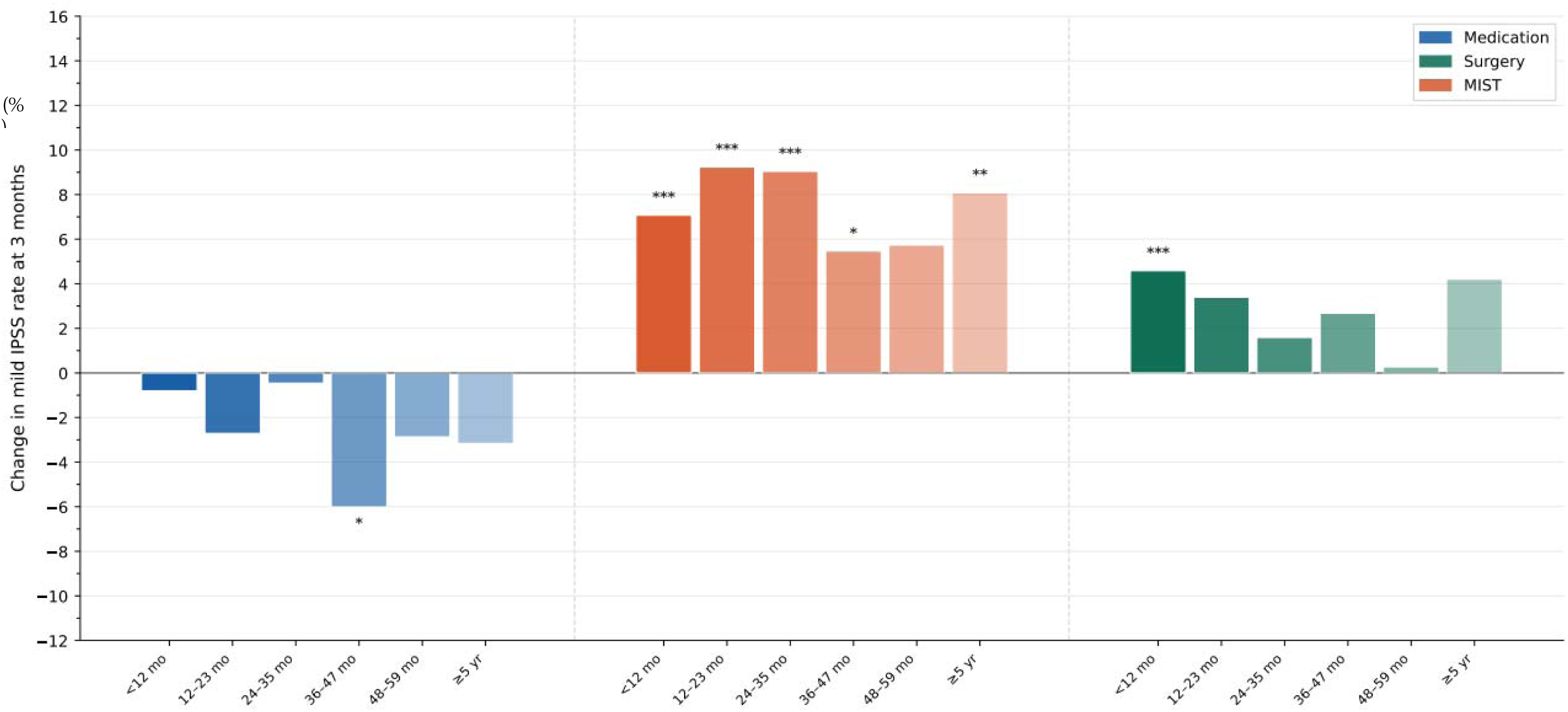
Percentage change in mild LUTS after 3 months from initiating treatment for BPH stratified by time from diagnosis to treatment. Asterisks denote significant difference between baseline and 3-month IPSS score at \**p* < 0.05, \*\**p* < 0.01, and \*\*\**p* < 0.001.

## RESULTS

### Baseline Characteristics

43,919 patients met criteria and were included in our analysis with baseline demographics summarized in Table 1. Patient care was provided by 4,060 urologists across 351 practices. Median age was 67 (IQR 61 – 73) with 13,586 patients (30.9%) between 70 and 80 years old and 4,022 (9.2%) > 80 years old. Median baseline IPSS was 10 (6 – 17) with most patients with baseline moderate symptoms (IPSS score 8 – 19; 24,911, 56.7%), though there were significant differences (*p* < 0.001) in baseline IPSS by index treatment, with surgery (15) and MIST (16) patients having a higher baseline IPSS. Comorbidities with significant differences (*p* < 0.001) included: diabetes (7.1%) and hypertension (17.4%) more common among the medical cohort, incontinence (9.8%) more common among the MIST cohort, and retention (28.9%) more common among the surgery cohort. One-third of the cohort (14,814 patients; 33.7%) had >5 years of follow-up available.

**Table 1.**
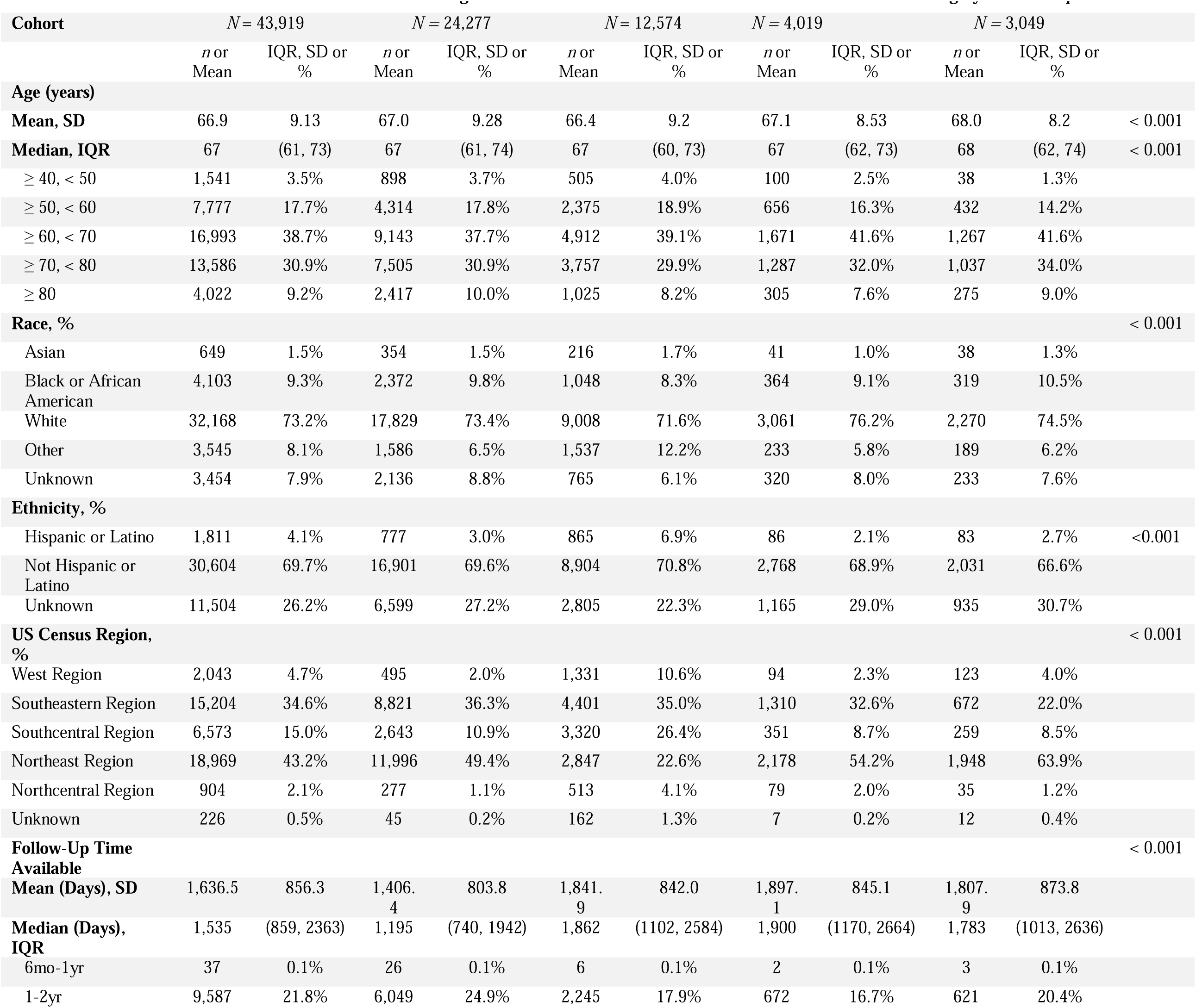
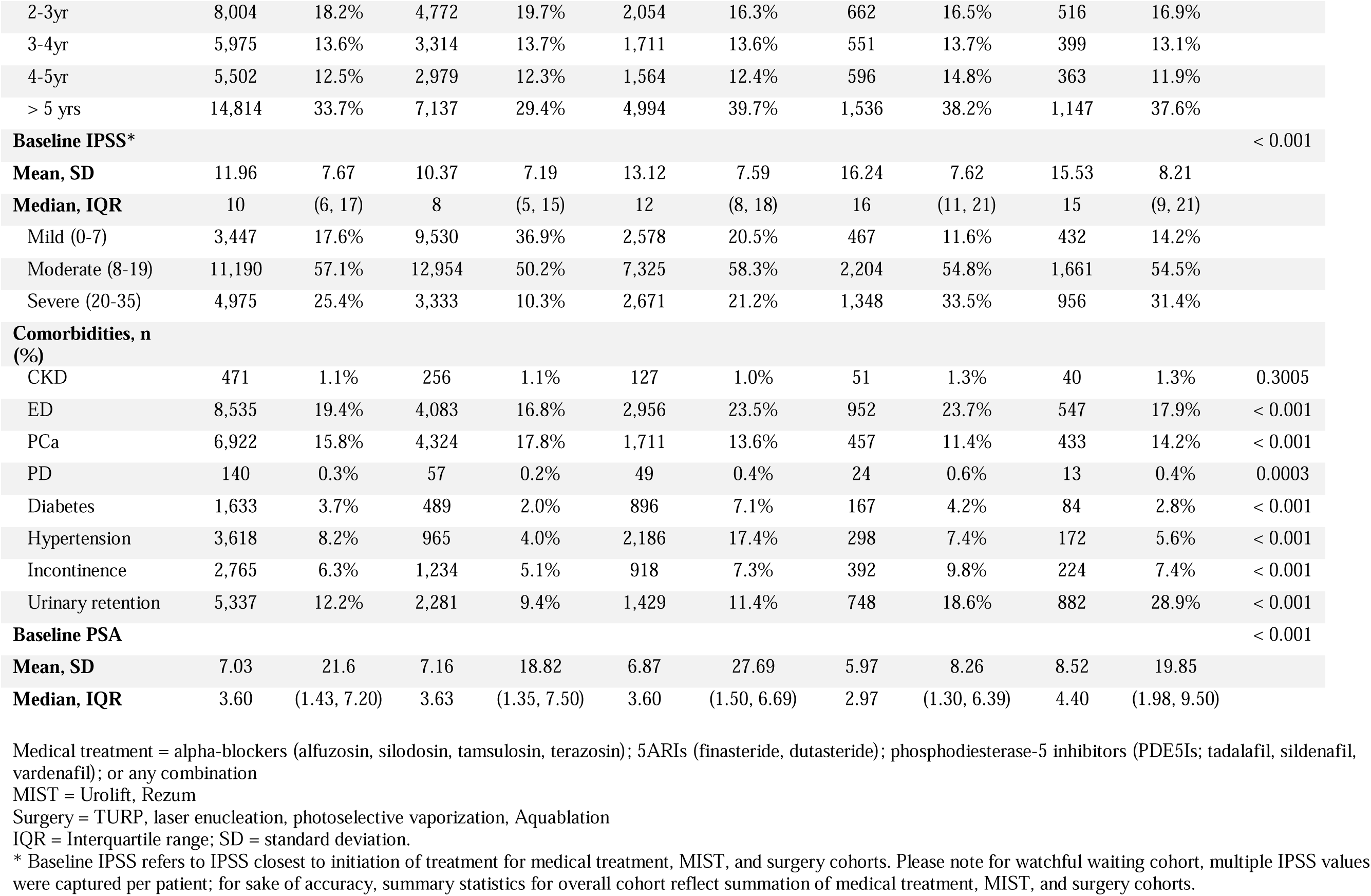
Baseline sociodemographic and clinical covariates stratified by treatment group.

### Change in IPSS Over Follow-Up

19,642 patients (44.7% of cohort) received some form of treatment within the study period. 12,574 patients (64.0%) pursued index medical treatment. Of those with multiple IPSS scores prior to starting therapy (42.4%; see Supplementary Table 1a), the median baseline IPSS score was 8 (7 – 15). Median IPSS at time of treatment was 12 (8 – 18) with 58.3% with moderate symptoms. Patients waited on average of 16.95 months before initiating treatment. Median IPSS after initiation of medical treatment was 12 (8 – 19) at 3 months, 11 (8 – 18) at 6 months, 10 (7 – 16) at 1 year, 10 (7– 16) at 2 years, and 10 (7 – 15) at 3 years follow-up. Patients who started medical treatment within 12 months of diagnosis reported significantly higher IPSS scores consistently across follow-up compared to patients who initiated therapy >12 months from diagnosis (*p* < 0.001; Figure 1a). Patients who initiated medical treatment <12 months from diagnosis had statistically significant improvements in IPSS scores out to 2 years of follow-up, while those who started therapy >12 months had less sustained improvements (Supplementary Table 2d).

4,019 patients (20.5%) underwent MIST as index treatment. Of those with multiple IPSS scores prior to MIST (86.3%; see Supplementary Table 1b), the median baseline IPSS was 16 (10 – 22). At time of MIST, median IPSS was 16 (11 – 21) with 54.8% with moderate symptoms. Patients waited an average of 24.05 months prior to pursuing MIST. Median IPSS after MIST was 14 (9 – 19) at 3 months, 13 (9 – 19) at 6 months, 12 (8 – 18) at 1 year, 12 (7 – 17) at 2 years, and 12 (7 – 17) at 3 years follow-up. Patients who pursued MIST within 12 months from diagnosis reported significantly higher IPSS scores consistently across follow-up compared to those who pursued MIST > 12 months from diagnosis (*p <* 0.001; Figure 1b). Patients who pursued MIST within 36 months of diagnosis generally had significant improvements in IPSS within 6-12 months of follow-up (*p* < 0.001; Supplementary Table 2l).

3,049 patients (15.5%) received surgical treatment as index treatment. Of those with multiple IPSS scores prior to surgery (87.2%; see Supplementary Table 1c), the median baseline IPSS was 14 (8 – 21). At time of surgery, median IPSS of 15 (9 – 21) with 54.5% with moderate symptoms. Patients waited on average 20.43 months before pursuing surgery. Median IPSS after surgical treatment was 14 (8 – 20) at 3 months, 12 (7 – 18) at 6 months, 11 (5 – 17) at 1 year, 11 (4 – 17) at 2 years, and 11 (4 – 17) at 3 years follow-up. There were no clinically meaningful differences in IPSS across follow-up as function of TTT (Figure 1c; significant differences found at 3 months and 1 year of follow-up were driven by small and clinically irrelevant differences). Patients who pursued surgery <5 years from diagnosis had significant improvements in IPSS within 6-12 months of follow-up, which was not observed among patients who delayed surgery to >5 years from treatment (*p* < 0.001; Supplementary Table 2h).

### Early Symptom Improvement

Patients who pursued index medical treatment for BPH exhibited no early symptom improvement as a function of TTT. By comparison, patients who pursued index surgical treatment exhibited early symptom improvement if they pursued surgery within 12 months of diagnosis (*p* < 0.001). Patients who pursued index MIST exhibited early symptom improvement regardless of when MIST was initiated from time of diagnosis (Figure 2; Supplementary Tables 3a – c; *p* < 0.01 to < 0.001).

### Procedural Events

Patients who started medical treatment within 12 months of diagnosis had a significantly longer time to event compared to those who delayed treatment >12 months from diagnosis (*p* < 0.001; Figure 3a). Patients who pursued MIST within 3 years of diagnosis had a significantly longer time to event than those who delayed treatment >3 years from diagnosis (*p* < 0.001; Figure 3b). Patients who sought surgery within 12 months of diagnosis had a significantly longer time to event compared to those who delayed treatment > 3 years from diagnosis (*p* < 0.001; Figure 3c; Supplementary Tables 4a – d).

**Figure 3.**
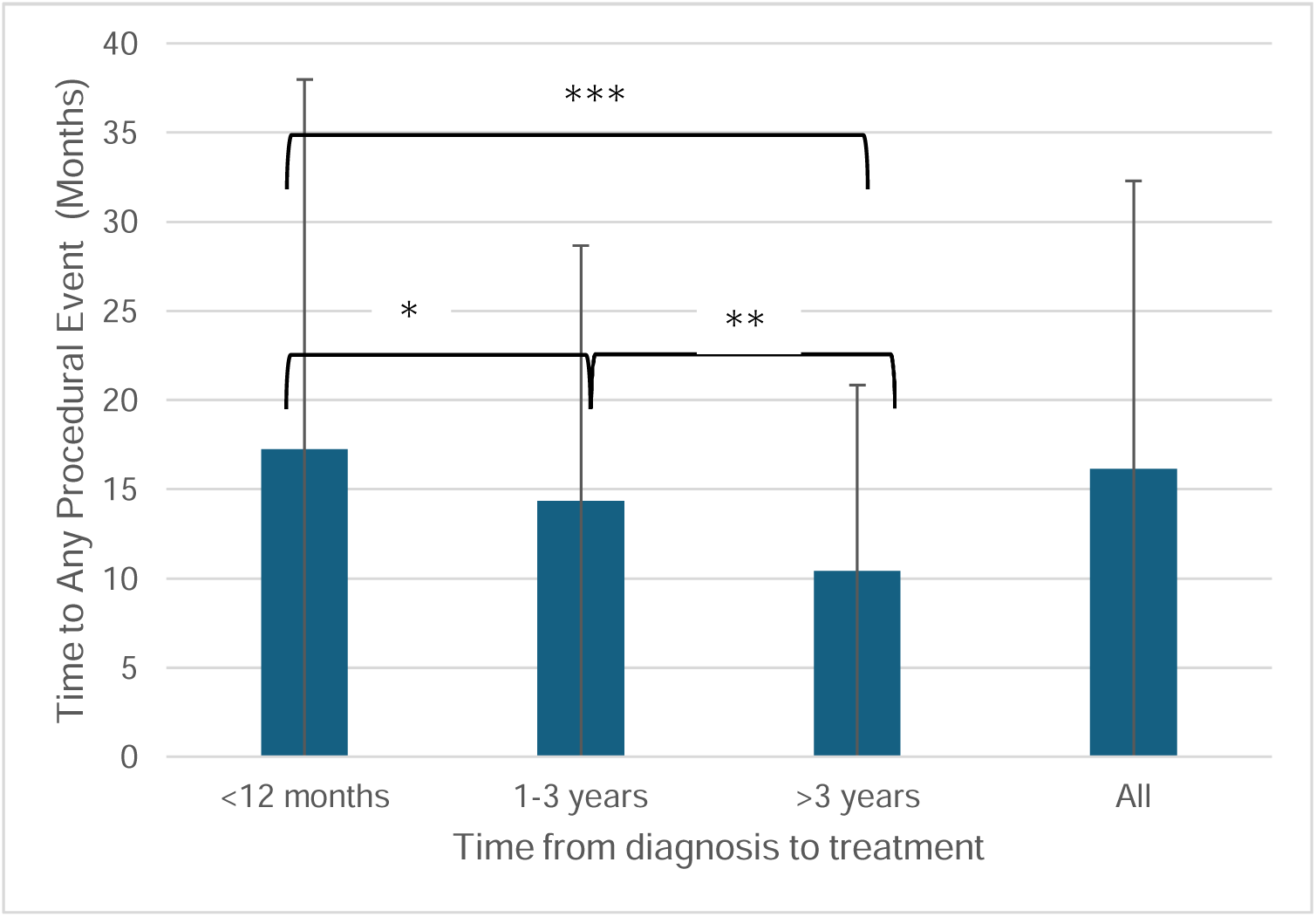
Time to any procedural event stratified by time from diagnosis to medical treatment for BPH. *denotes *p* < 0.05, ** *p* < 0.01, *** *p* < 0.001.

**Figure 3b.**
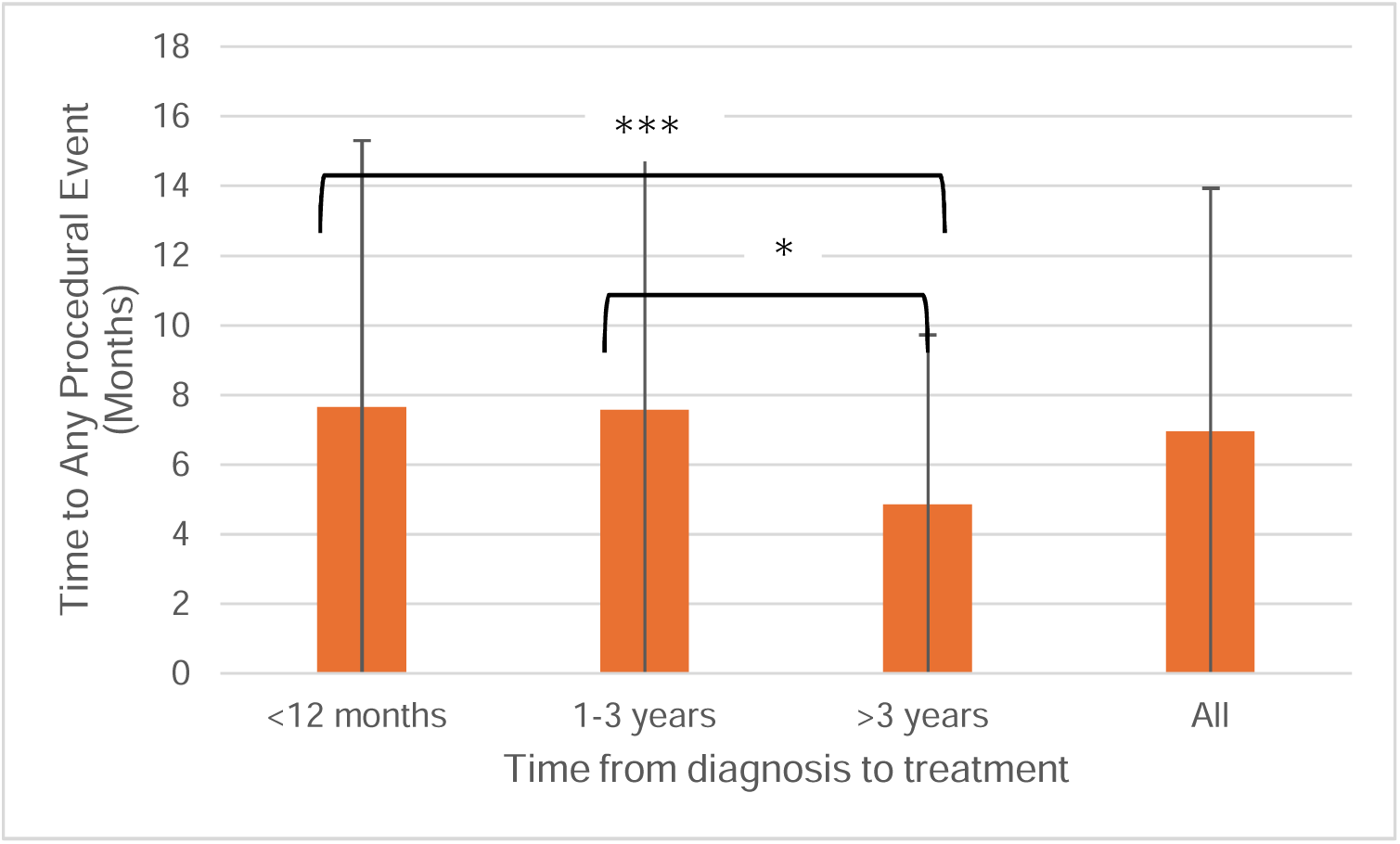
Time to any procedural event stratified by time from diagnosis to MIST for BPH. *denotes *p* < 0.05, *p* < 0.001.

**Figure 3c.**
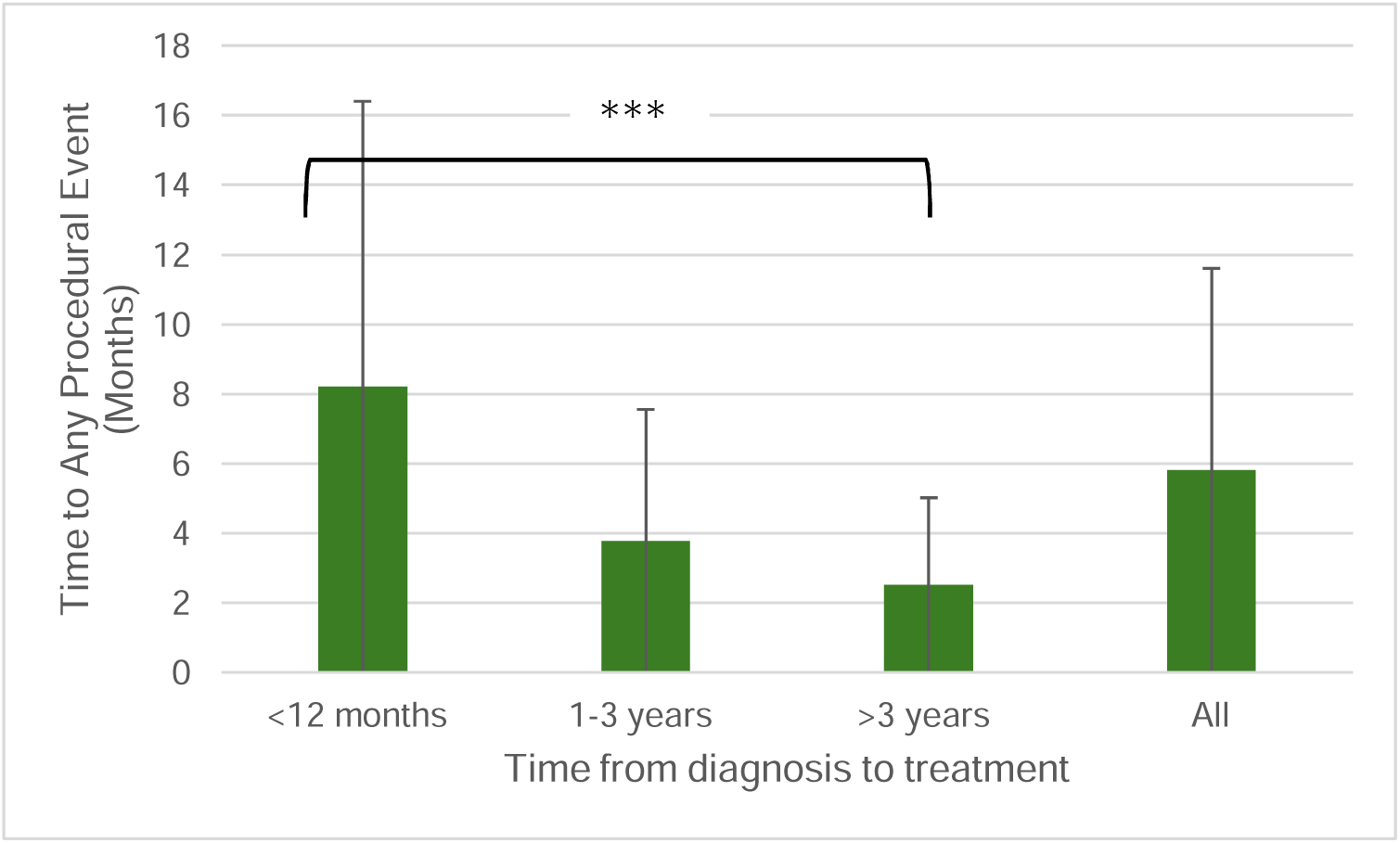
Time to any procedural event stratified by time from diagnosis to surgical treatment for BPH. *** denotes *p* < 0.001.

### Time to Retreatment

Mean time to retreatment for patients who pursued medical treatment initially or surgical re-treatment for patients who pursued MIST or surgery initially are included in Figure 4. Patients who pursued medical treatment within 3 years of BPH diagnosis had a significantly longer time to retreatment, including initiation of OAB medications (*p* < 0.001) and pursuing surgery or MIST (*p* < 0.01), though no significant patterns were observed for patients undergoing index surgical re-treatment after pursuing index MIST or surgery (Supplementary Tables 5a – f).

**Figure 4.**
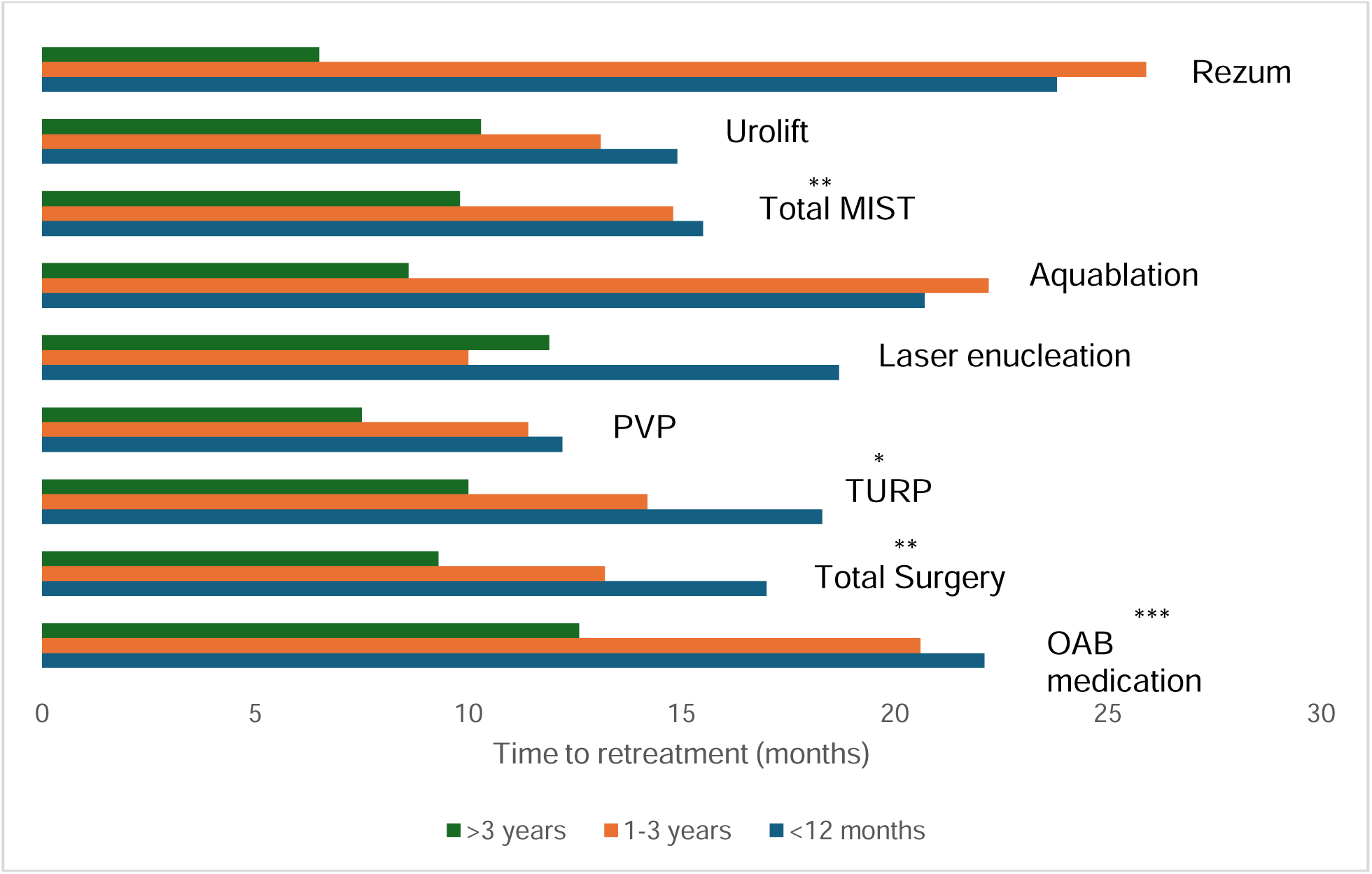
Time to secondary retreatment after index medical treatment for BPH stratified by TTT. OAB medication refers to addition of new OAB medication. Total surgery includes TURP, PVP, laser enucleation, and Aquablation. Total MIST includes Urolift and Rezum. * denotes *p* < 0.5, ** denotes *p* < 0.01, *** denotes *p* < 0.001; statistical comparison refers to overall differences with each category.

## DISCUSSION

We present a novel analysis on the impact of timing of BPH treatment on symptom improvement among a contemporary cohort. Our findings highlight a benefit to pursuing treatment sooner on delaying subsequent procedural events, and, in the case of the surgical cohorts, early symptom improvement. Our study has important strengths; use of the AQUA registry allows for an important analysis of contemporary practice patterns which are otherwise challenging to assess at this scale for BPH therapies. We also formally assess the impact of time-to-treatment on outcomes, which we believe is particularly relevant to maximize patient satisfaction and minimize potential regret when pursuing elective BPH treatment.

We observed a median change in IPSS score of 1.0 at 6 months of follow-up among men on pharmacotherapy within 12 months of diagnosis, which is less than the accepted minimal clinically significant difference of 3.^17^ Alcaraz, *et al.* reported a mean change in IPSS of 2.4 and 2.8 for monotherapy and combination therapy, respectively, among men with similar baseline IPSS.^18^ Long-term IPSS changes among our cohort were similarly less pronounced than reported in landmark studies, though median baseline IPSS scores for these studies were approximately 15 – 17 compared to median 8 – 13 herein.^3–4^ Both procedural cohorts were also less symptomatic at baseline and reported less pronounced improvement in LUTS over follow-up compared to relevant prospective trials. ^8–10, 19–20^ Our findings may highlight a floor effect; that is, men with lower baseline LUTS may report less of a noticeable effect in their symptoms. Kaplan, *et al.* demonstrated that men with “low-moderate” baseline LUTS (e.g. IPSS 8-12) reported less than a 2-point reduction in IPSS score after 4 years of finasteride therapy.^21^ This effect has also been demonstrated in a subgroup analysis of the WATER trial.^22^ Given that men are pursuing treatment for relatively modest LUTS, it is possible that these decisions are being driven by bother.

Men who delayed surgery >5 years or MIST > 36 months from time of diagnosis did not exhibit the significant decline in IPSS within 6-12 months of surgery observed in those who pursued surgery sooner. These data suggest that surgery and MIST may afford a similar benefit to patients until it is delayed to a point at which disease progression overwhelms benefit. Bladder cellular degeneration has been correlated with age, and these ultrastructural detrusor muscle changes are associated with voiding dysfunction after TURP.^23^ Pyun, *et al.* demonstrated that men with evidence of detrusor over- or underactivity on preoperative urodynamics demonstrated poorer postoperative IPSS scores compared to men with outlet obstruction without preoperative detrusor changes.^24^ While we do not have data on bother scores alone, it is possible that men who delayed intervention rated higher bother scores postoperatively reflecting their experience with complications or ongoing urinary symptoms.

Our cohort had a high rate of index MIST and surgery at 20.5% and 15.5%, respectively. We suspect this may be related to the large urology group practices that are enrolled in the AQUA registry. These practices are likely to have specialized urologists who have experience in MIST and surgical therapies; their comfort level with offering upfront procedural intervention may be higher, with prior patient experience and outcomes informing their index treatment offerings. Procedural intervention is likely attractive to many patients as well who prefer this approach over the prospect of lifelong medication and side effects.

While there was no significant increase in mild LUTS within 3 months of initiating pharmacotherapy regardless of when men pursued treatment, there was a significant increase in mild LUTS among patients who pursued surgery within 12 months of diagnosis and among all MIST patients regardless of TTT. These findings in part reflect a ceiling effect; that is, given that there was a higher proportion of mild LUTS at baseline among the medicine cohort compared to the procedural cohorts, a greater increase in mild LUTS among this cohort would be required to achieve significance. However, it remains important to note that despite the benefits of treatment on voiding parameters,^3,4^ changes to voiding parameters alone do not necessarily correlate with improvements in patient-reported benefit.^25^

Decisional regret after BPH treatment may in part inform patient-reported benefit. In a prospective cohort of men undergoing surgery or MISTs, Winograd *et al.* demonstrated that one-third of their cohort reported significant regret with pursuing treatment, with most common reasons for regret including perceived lack of efficacy, new urinary symptoms, and postoperative complications. Importantly, they found that patients with higher regret reported significantly reduced change in IPSS with surgical treatment.^26^ Regret may also be driven by poorer patient selection, as demonstrated among patients with larger gland size undergoing MIST.^27^ Given that the median baseline IPSS scores were highest across all TTT cohorts pursuing MIST and the adverse event profile is generally more favorable for MIST compared to surgery,^11–12^ it is possible that the significant increase in mild LUTS across all TTT cohorts reflects favorable conditions associated with lower decisional regret. Similarly, lack of early improvement across all TTT groups pursuing medical treatment may reflect patient dissatisfaction with perceived time-to-effect, bother with associated side effects,^28^ and incongruence between physician and patient goals of treatment.

Early treatment in our cohort was significantly associated with delayed time of procedural events. Among the medical cohort, these findings may reflect increased prescribing of dual pharmacotherapy with 5ARIs,^29^ which have been established to reduce likelihood of urinary retention and progression to surgery.^3,4^ Delaying surgical treatment may predispose patients to changes in detrusor function or larger gland size that ultimately led to further disease progression. Importantly, given that events on average occurred well beyond 30 days of intervention, we are assured that they do not reflect early postoperative complications.

There are several important limitations to note. We found significant differences in modality of index treatment by geographic region; these differences may reflect differences in access to care and reimbursement patterns for participating practices. Additionally, differences in health literacy between these populations may impact patient-reported outcomes. Furthermore, while all patients in our study had available IPSS data, 85.5% of patients in the BPH registry do not have available IPSS scores. This was a shocking finding as IPSS is consistently used across all studies to measure disease progression and treatment response. The AUA guidelines for BPH also recommend obtaining a baseline IPSS score and repeating at 4-12 weeks follow-up after initiating treatment.^30^ It is possible that IPSS is assessed and not formally recorded. Regardless, as the AQUA Registry allows providers to be compensated for reporting various urology-specific quality metrics in their practice as part of a Merit-Based Incentive Payment System, this does show a needed area of improvement. Additionally, patients with available IPSS data were younger, more likely to pursue MIST and surgery, and more likely to receive care in the Northeast (see Supplementary Table 6). Lastly, while statistically significant differences in IPSS may not reflect clinically meaningful differences, they do allow for comparison between TTT cohorts.

## CONCLUSIONS

Among a contemporary cohort reflecting real-world BPH practice using the AQUA Registry, we found that pursuing procedural intervention sooner from time of diagnosis was associated with significant symptom improvement. Early BPH treatment was also associated with delayed time to procedural events. With patients pursuing higher rates of index surgery for generally modest LUTS, providers should establish patient goals and set expectations early to minimize potential regret.

## Data Availability

Data are available from the authors upon reasonable request and with permission of Verana, though which data were licensed.

## ABBREVIATIONS

5ARI: 5α-reductase inhibitor
AQUA: American Urological Association Quality Registry
BPH: Benign prostatic hyperplasia
IPSS: International Prostate Symptom Score
LUTS: Lower urinary tract symptoms
OAB: Overactive bladder
MIST: Minimally invasive surgical therapy
PDE5I: Phosphodiesterase-5 inhibitor
PVP: Photoselective vaporization of prostate
TTT: Time-to-treatment
TURP: Transurethral resection of prostate

## ACKNOWLEDGEMENTS

The authors have no acknowledgements.

