## Supplementary Tables for "Impact of Early Treatment on Symptom Improvement and Procedural Events among Men with BPH and Bothersome Lower Urinary Tract Symptoms: A Contemporary Analysis of the American Urological Association Quality (AQUA) Registry"

SUPPLEMENTARY TABLES AND FIGURES

Table 1a. Changes in IPSS scores for patients pursuing medical treatment for BPH stratified by time-to-treatment.

Table 1b. Change in IPSS scores for patients pursuing surgical treatment for BPH stratified by time-to-treatment.

Table 1c. Change in IPSS scores for patients pursuing MIST for BPH stratified by time-to-treatment.

Table 2a. Comparison of median IPSS scores between TTT groups undergoing medical treatment over follow-up.

Table 2b. Pairwise comparison of median IPSS scores between TTT groups undergoing medical treatment over follow-up.

Table 2c. Comparison of median IPSS score over time within each TTT group pursuing medical treatment.

Table 2d. Pairwise comparison of median IPSS score over time within each TTT group pursuing medical treatment.

Table 2e. Comparison of median IPSS scores between TTT groups undergoing surgery over follow-up.

Table 2f. Pairwise comparison of median IPSS scores between TTT groups undergoing surgery over follow-up.

Table 2g. Comparison of median IPSS score over time within each TTT group pursuing surgery.

Table 2h. Pairwise comparison of median IPSS score over time within each within each TTT group pursuing surgery.

Table 2i. Comparison of median IPSS scores between TTT groups undergoing MIST over follow-up.

Table. 2j. Pairwise comparison of median IPSS scores between TTT groups undergoing MIST over follow-up.

Table 2k. Comparison of median IPSS score over time within each TTT group pursuing MIST.

Table 2l. Pairwise comparison of median IPSS score over time within each TTT group pursuing MIST.

Table 3a. Change from moderate-to-severe to mild LUTS after 3 months from initiating medical treatment for BPH stratified by time from diagnosis to treatment.

Table 3b. Change from moderate-to-severe to mild LUTS after 3 months from initiating surgical treatment for BPH stratified by time from diagnosis to treatment.

Table 3c. Change from moderate-to-severe to mild LUTS after 3 months from initiating MIST for BPH stratified by time from diagnosis to treatment.

Table 4a. Time-to-event stratified by time from diagnosis to medical treatment for BPH.

Table 4b. Time-to-event stratified by time from diagnosis to surgical treatment for BPH.

Table 4c. Time-to-event stratified by time from diagnosis to MIST for BPH.

Table 4d. Pairwise comparisons of mean time-to-event stratified by time from diagnosis to treatment.

Table 5a. Time to secondary retreatment after index medical treatment for BPH stratified by time from diagnosis to treatment.

Table 5b. Time to surgical retreatment after index surgical treatment for BPH stratified by time from diagnosis to treatment.

Table 5c. Time to surgical retreatment after index MIST treatment for BPH stratified by time from diagnosis to treatment.

Table 5d. Pairwise comparisons of time to secondary treatment after index medical treatment for BPH by time from diagnosis to treatment.

Table 5e. Pairwise comparisons of time to secondary treatment after index surgical treatment for BPH by time from diagnosis to treatment.

Table 5f. Pairwise comparisons of time to secondary treatment after index MIST for BPH by time from diagnosis to treatment.

Table 6. Clinicodemographic variables for patients with and without available IPSS scores within AQUA Registry.

Table 1a. Changes in IPSS scores for patients pursuing medical treatment for BPH stratified by time-to-treatment.

|  | *<12 months from diagnosis*  *N =7,726* | | *12-23 months from diagnosis*  *N = 1,449* | | *24-35 months from diagnosis*  *N = 946* | | *36-47 months from diagnosis*  *N = 747* | | *48-59 months from diagnosis*  *N = 590* | | *>60 months from diagnosis*  *N = 1,116* | | *All patients*  *N = 12,574* | |
| --- | --- | --- | --- | --- | --- | --- | --- | --- | --- | --- | --- | --- | --- | --- |
|  | ***n*** | **SD/IQR or %** | ***n*** | **SD/IQR or %** | ***n*** | **SD/IQR or %** | ***n*** | **SD/IQR or %** | ***n*** | **SD/IQR or %** | ***n*** | **SD/IQR or %** | ***n*** | **SD/IQR or %** |
| Mean number of IPSS evaluations between diagnosis and treatment | 1.49 | 1.41 | 3.16 | 3.32 | 3.86 | 4.34 | 4.07 | 4.91 | 4.25 | 4.60 | 5.20 | 6.28 | 2.47 | 3.40 |
| Mean time between diagnosis and treatment (months) | 1.66 | 2.97 | 16.9 | 3.66 | 29.39 | 3.46 | 41.67 | 3.62 | 53.42 | 3.44 | 76.41 | 11.70 | 16.95 | 24.27 |
| *Before treatment* |  |  |  |  |  |  |  |  |  |  |  |  |  |  |
| First pre-treatment IPSS | 2,087 | 27.01% | 977 | 67.43% | 654 | 69.13% | 490 | 65.60% | 376 | 63.73% | 748 | 67.03% | 5,332 | 42.41% |
| Mean | 12.11 | 7.60 | 10.40 | 6.65 | 10.26 | 6.65 | 9.91 | 5.97 | 10.16 | 6.21 | 9.81 | 6.04 | 10.91 | 6.94 |
| Median | 10 | (8, 17) | 8 | (7, 13) | 8 | (7, 14) | 8 | (7, 13) | 8 | (7, 13) | 8 | (7, 12) | 8 | (7, 15) |
| Mild (0-7) | 507 | 24.29% | 268 | 27.43% | 185 | 28.29% | 133 | 27.14% | 102 | 27.13% | 197 | 26.34% | 1,392 | 26.11% |
| Moderate (8-19) | 1,188 | 56.92% | 608 | 62.23% | 396 | 60.55% | 320 | 65.31% | 237 | 63.03% | 490 | 65.51% | 3,239 | 60.75% |
| Severe (20-35) | 392 | 18.78% | 101 | 10.34% | 73 | 11.16% | 37 | 7.55% | 37 | 9.84% | 61 | 8.16% | 701 | 13.15% |
| Last pre-treatment IPSS | 7,726 | 100.00% | 1,449 | 100.00% | 946 | 100.00% | 747 | 100.00% | 590 | 100.00% | 1,116 | 100.00% | 12,574 | 100.00% |
| Mean | 14.24 | 7.76 | 11.49 | 7.06 | 11.20 | 6.9 | 11.36 | 6.78 | 11.72 | 7.10 | 11.00 | 6.7 | 13.12 | 7.59 |
| Median | 13 | (8, 20) | 10 | (8, 16) | 8 | (7, 16) | 9 | (8, 15) | 10 | (8, 16) | 8 | (8, 15) | 12 | (8, 18) |
| Mild (0-7) | 1,431 | 18.52% | 343 | 23.67% | 242 | 25.58% | 172 | 23.03% | 130 | 22.03% | 260 | 23.30% | 2,578 | 20.50% |
| Moderate (8-19) | 4,274 | 55.32% | 898 | 61.97% | 582 | 61.52% | 480 | 64.26% | 371 | 62.88% | 720 | 64.52% | 7,325 | 58.26% |
| Severe (20-35) | 2,021 | 26.16% | 208 | 14.35% | 122 | 12.90% | 95 | 12.72% | 89 | 15.08% | 136 | 12.19% | 2,671 | 21.24% |
| *3 months post-treatment* | |  |  |  |  |  |  |  |  |  |  |  |  |  |
| 3-month follow-up IPSS | 4,120 | 53.33% | 606 | 41.82% | 414 | 43.76% | 335 | 44.85% | 287 | 48.64% | 581 | 52.06% | 6,343 | 50.45% |
| Mean | 14.45 | 7.79 | 12.16 | 7.4 | 11.69 | 7.23 | 12.00 | 6.85 | 12.04 | 7.32 | 11.31 | 6.55 | 13.53 | 7.66 |
| Median | 14 | (8, 20) | 10 | (8, 17) | 9 | (7, 17) | 10 | (8, 16) | 10 | (8, 15) | 9 | (8, 15) | 12 | (8, 19) |
| Mild (0-7) | 730 | 17.72% | 127 | 20.96% | 104 | 25.12% | 57 | 17.01% | 55 | 19.16% | 117 | 20.14% | 1,190 | 18.76% |
| Moderate (8-19) | 2,293 | 55.66% | 376 | 62.05% | 242 | 58.45% | 229 | 68.36% | 185 | 64.46% | 391 | 67.30% | 3,716 | 58.58% |
| Severe (20-35) | 1,097 | 26.63% | 103 | 17.00% | 68 | 16.43% | 49 | 14.63% | 47 | 16.38% | 73 | 12.56% | 1,437 | 22.65% |
| Change from baseline IPSS |  |  |  |  |  |  |  |  |  |  |  |  |  |  |
| Mean, SD | -0.75 | 4.49 | -0.40 | 4.41 | -0.41 | 3.65 | -0.16 | 3.37 | -0.32 | 3.93 | -0.26 | 3.80 | -0.60 | 4.30 |
| Median, IQR | 0 | (0, 0) | 0 | (0, 0) | 0 | (0, 0) | 0 | (0, 0) | 0 | (0, 0) | 0 | (0, 0) | 0 | (0, 0) |
| Clinic visits per patient | 7,726 | 100.00% | 1,449 | 100.00% | 946 | 100.00% | 747 | 100.00% | 590 | 100.00% | 1,116 | 100.00% | 12,574 | 100.00% |
| Mean, SD | 1.64 | 2.4 | 1.17 | 2.47 | 1.19 | 2.16 | 1.15 | 2.22 | 1.47 | 3.49 | 1.26 | 2.59 | 1.48 | 2.48 |
| Median, IQR | 1 | (0, 2) | 1 | (0, 1) | 1 | (0, 2) | 1 | (0, 1) | 1 | (0, 2) | 1 | (0, 2) | 1 | (0, 2) |
| *6 months post-treatment* | |  |  |  |  |  |  |  |  |  |  |  |  |  |
| 6-month follow-up IPSS | 2,523 | 32.66% | 432 | 29.81% | 275 | 29.07% | 227 | 30.39% | 192 | 32.54% | 343 | 30.73% | 3,992 | 31.75% |
| Mean | 13.63 | 7.77 | 11.08 | 6.52 | 10.86 | 6.49 | 11.09 | 6.73 | 11.61 | 6.75 | 11.07 | 6.79 | 12.70 | 7.47 |
| Median | 13 | (8, 19) | 8 | (8, 15) | 8 | (7, 15) | 10 | (7, 15) | 10 | (8, 15) | 8 | (7, 15) | 11 | (8, 18) |
| Mild (0-7) | 521 | 20.65% | 106 | 24.54% | 76 | 27.64% | 60 | 26.43% | 37 | 19.27% | 86 | 25.07% | 886 | 22.19% |
| Moderate (8-19) | 1,417 | 56.16% | 277 | 64.12% | 165 | 60.00% | 138 | 60.79% | 129 | 67.19% | 207 | 60.35% | 2,333 | 58.44% |
| Severe (20-35) | 585 | 23.19% | 49 | 11.34% | 34 | 12.36% | 29 | 12.78% | 26 | 13.54% | 50 | 14.58% | 773 | 19.36% |
| Change from baseline IPSS |  |  |  |  |  |  |  |  |  |  |  |  |  |  |
| Mean, SD | -1.29 | 5.27 | -0.70 | 4.31 | -0.36 | 4.64 | -1.09 | 4.56 | -0.64 | 4.64 | -0.27 | 3.35 | -1.03 | 4.93 |
| Median, IQR | 0 | ((0, 0) | 0 | ((0, 0) | 0 | (0, 0) | 0 | (0, 0) | 0 | (0, 0) | 0 | (0, 0) | 0 | (0, 0) |
| Clinic visits per patient | 7,726 | 100.00% | 1,449 | 100.00% | 946 | 100.00% | 747 | 100.00% | 590 | 100.00% | 1,116 | 100.00% | 12,574 | 100.00% |
| Mean, SD | 0.9 | 1.63 | 0.7 | 1.36 | 0.75 | 1.66 | 0.75 | 1.37 | 0.86 | 1.54 | 0.82 | 2.47 | 0.89 | 1.68 |
| Median, IQR | 0 | (0, 1) | 0 | (0, 1) | 0 | (0, 1) | 0 | (0, 1) | 0 | (0, 1) | 0 | (0, 1) | 0 | (0, 1) |
| *1-year post-treatment* | |  |  |  |  |  |  |  |  |  |  |  |  |  |
| 1-year follow-up IPSS | 3,690 | 47.76% | 696 | 48.03% | 434 | 45.88% | 341 | 45.65% | 251 | 42.54% | 430 | 38.53% | 5,842 | 46.46% |
| Mea | 12.43 | 7.37 | 10.59 | 6.62 | 10.52 | 6.54 | 10.80 | 6.82 | 10.92 | 6.34 | 10.34 | 6.08 | 11.76 | 7.12 |
| Median | 11 | (8, 17) | 8 | (7, 14) | 8 | (7, 14) | 9 | (6, 15) | 10 | (8, 14) | 8 | (8, 14) | 10 | (7, 16) |
| Mild (0-7) | 913 | 24.74% | 197 | 28.30% | 126 | 29.03% | 98 | 28.74% | 61 | 24.30% | 107 | 24.88% | 1,502 | 25.71% |
| Moderate (8-19) | 2,137 | 57.91% | 430 | 61.78% | 257 | 59.22% | 206 | 60.41% | 162 | 64.54% | 289 | 67.21% | 3,481 | 59.59% |
| Severe (20-35) | 640 | 17.34% | 69 | 9.91% | 51 | 11.75% | 37 | 10.85% | 28 | 11.16% | 34 | 7.91% | 859 | 14.70% |
| Change from baseline IPSS |  |  |  |  |  |  |  |  |  |  |  |  |  |  |
| Mean, SD | -1.59 | 5.49 | -0.58 | 4.72 | -0.36 | 4.70 | -0.99 | 4.87 | -0.63 | 4.19 | -0.61 | 4.36 | -1.23 | 5.20 |
| Median, IQR | 0 | (-2, 0) | 0 | (0, 0) | 0 | (0, 0) | 0 | (0, 0) | 0 | (0, 0) | 0 | (0, 0) | 0 | (-1, 0) |
| Clinic visits per patient | 7,726 | 100.00% | 1,449 | 100.00% | 946 | 100.00% | 747 | 100.00% | 590 | 100.00% | 1,116 | 100.00% | 12,574 | 100.00% |
| Mean, SD | 1.70 | 2.79 | 1.38 | 1.92 | 1.24 | 1.95 | 1.35 | 2.04 | 1.31 | 1.96 | 1.00 | 1.98 | 1.53 | 2.52 |
| Median, IQR | 1 | (0, 2) | 1 | (0, 2) | 1 | (0, 2) | 1 | (0, 2) | 1 | (0, 2) | 0 | (0, 1) | 1 | (0, 2) |
| *2-year post-treatment* | |  |  |  |  |  |  |  |  |  |  |  |  |  |
| 2-year follow-up IPSS | 4,405 | 57.02% | 694 | 47.90% | 447 | 47.25% | 351 | 46.99% | 223 | 37.80% | 374 | 33.51% | 6,494 | 51.65% |
| Mean | 12.22 | 7.29 | 10.11 | 6.4 | 9.63 | 6.10 | 10.31 | 6.58 | 11.15 | 6.86 | 9.94 | 6.25 | 11.54 | 7.09 |
| Median | 11 | (7, 17) | 8 | (6. 14) | 8 | (5, 13) | 8 | (6, 14) | 10 | (7, 15) | 9 | (6, 13) | 10 | (7, 16) |
| Mild (0-7) | 1,112 | 25.24% | 210 | 30.26% | 143 | 31.99% | 104 | 29.63% | 56 | 25.11% | 109 | 29.14% | 1,734 | 26.70% |
| Moderate (8-19) | 2,594 | 58.89% | 421 | 60.66% | 265 | 59.28% | 214 | 60.97% | 142 | 63.68% | 233 | 62.30% | 3,869 | 59.58% |
| Severe (20-35) | 699 | 15.87% | 63 | 9.08% | 39 | 8.72% | 33 | 9.40% | 25 | 11.21% | 32 | 8.56% | 891 | 13.72% |
| Change from baseline IPSS |  |  |  |  |  |  |  |  |  |  |  |  |  |  |
| Mean, SD | -1.84 | 5.97 | -0.69 | 4.98 | -0.58 | 5.50 | -0.69 | 5.40 | -0.04 | 5.44 | -0.57 | 4.92 | -1.43 | 5.76 |
| Median, IQR | 0 | (-3, 0) | 0 | (-1, 0) | 0 | (0, 0. | 0 | (-1, 0) | 0 | (0, 0) | 0 | (-1, 0) | 0 | (-2, 0) |
| Clinic visits per patient | 7,726 | 100.00% | 1,449 | 100.00% | 946 | 100.00% | 747 | 100.00% | 590 | 100.00% | 1,116 | 100.00% | 12,574 | 100.00% |
| Mean, SD | 2.50 | 3.24 | 1.81 | 2.86 | 1.73 | 3.09 | 1.91 | 3.59 | 1.52 | 2.54 | 1.0 | 2.05 | 2.16 | 3.12 |
| Median, IQR | 2 | (1, 3) | 1 | (0, 2) | 1 | (0, 2) | 1 | (0, 2) | 1 | (0, 2) | 0 | (0, 1) | 1 | (0, 3) |
| *3-year post-treatment* | |  |  |  |  |  |  |  |  |  |  |  |  |  |
| 3-year follow-up IPSS | 3,762 | 48.69% | 645 | 44.51% | 372 | 39.32% | 262 | 35.07% | 151 | 25.59% | 152 | 13.62% | 5,344 | 42.50% |
| Mean | 11.83 | 7.17 | 10.07 | 6.07 | 9.86 | 5.96 | 10.29 | 6.31 | 9.3 | 6.06 | 10.32 | 5.91 | 11.29 | 6.92 |
| Median | 11 | (7, 16) | 8 | (7, 13) | 8 | (7, 13) | 8 | (7, 14) | 8 | (5, 12) | 10 | (7, 14) | 10 | (7, 15) |
| Mild (0-7) | 991 | 26.34% | 180 | 27.91% | 106 | 28.49% | 70 | 26.72% | 51 | 33.77% | 41 | 26.97% | 1,439 | 26.93% |
| Moderate (8-19) | 2,207 | 58.67% | 409 | 63.41% | 236 | 63.44% | 170 | 64.89% | 92 | 60.93% | 103 | 67.76% | 3,217 | 60.20% |
| Severe (20-35) | 564 | 14.99% | 56 | 8.68% | 30 | 8.06% | 22 | 8.40% | 8 | 5.30% | 8 | 5.26% | 688 | 12.87% |
| Change from baseline IPSS |  |  |  |  |  |  |  |  |  |  |  |  |  |  |
| Mean, SD | -1.88 | 6.39 | -0.7 | 5.38 | -0.41 | 5.31 | 0.03 | 4.67 | -0.73 | 5.06 | -0.51 | 5.33 | -1.47 | 6.10 |
| Median, IQR | 0 | (-4, 0) | 0 | (-1, 0) | 0 | (-1. 0) | 0 | (-1, 0) | 0 | (-2, 0. | 0 | (-1, 0) | 0 | (-3, 0) |
| Clinic visits per patient | 7,726 | 100.00% | 1,449 | 100.00% | 946 | 100.00% | 747 | 100.00% | 590 | 100.00% | 1,116 | 100.00% | 12,574 | 100.00% |
| Mean, SD | 5.0 | 7.8 | 3.73 | 6.07 | 3.06 | 5.40 | 2.53 | 5.29 | 1.38 | 4.14 | 0.49 | 1.53 | 4.00 | 6.98 |
| Median, IQR | 3 | (0, 7) | 1 | (0, 5) | 1 | (0, 4) | 0 | (0, 3) | 0 | (0, 1) | 0 | (0, 0) | 2 | (0, 5) |

IQR = Interquartile range; SD = standard deviation; IPSS = International Prostate Symptom Score.

Table 1b. Change in IPSS scores for patients pursuing surgical treatment for BPH stratified by time-to-treatment.

|  | | *<12 months from diagnosis*  *N = 1,533* | | | *12-23 months from diagnosis*  *N = 541* | | *24-35 months from diagnosis*  *N = 350* | | | *36-47 months from diagnosis*  *N = 215* | | | *48-59 months from diagnosis*  *N = 171* | | | *>60 months from diagnosis*  *N = 239* | | | *All patients*  *N = 3,049* | | |
| --- | --- | --- | --- | --- | --- | --- | --- | --- | --- | --- | --- | --- | --- | --- | --- | --- | --- | --- | --- | --- | --- |
|  | | ***n*** | | **SD/IQR or %** | ***n*** | **SD/IQR or %** | ***n*** | | **SD/IQR or %** | ***n*** | | **SD/IQR or %** | ***n*** | | **SD/IQR or %** | ***n*** | | **SD/IQR or %** | ***n*** | **SD/IQR or %** | |
| Mean number of IPSS evaluations between diagnosis and treatment | 3.23 | | 2.03 | | 4.7505 | 2.88 | | 5.56 | 3.36 | | 5.59 | 3.76 | | 6.16 | 4.53 | | 7.08 | 6.22 | 4.40 | | 3.42 |
| Mean time between diagnosis and treatment (months) | 4.13 | | 3.16 | | 17.10 | 3.39 | | 29.43 | 3.24 | | 41.53 | 3.56 | | 53.41 | 3.52 | | 76.51 | 11.76 | 20.43 | | 22.46 |
| *Before treatment* | | | | | | | | | | | | | | | | | | | | | |
| First pre-treatment IPSS | | 1,277 | | 83.30% | 496 | 91.68% | 322 | | 92.00% | 194 | | 90.23% | 155 | | 90.64% | 214 | | 89.54% | 2,658 | 87.18% | |
| Mean | | 16.39 | | 8.38 | 14.33 | 7.74 | 14.89 | | 8.13 | 12.57 | | 8.09 | 14.44 | | 8.30 | 12.84 | | 7.84 | 15.15 | 8.27 | |
| Median | | 16 | | (9, 23) | 14 | (8, 20) | 14 | | (8, 20) | 11 | | (7, 17) | 13 | | (8, 20) | 12 | | (7, 19) | 14 | (8, 21) | |
| Mild (0-7) | | 153 | | 11.98% | 86 | 17.34% | 62 | | 19.25% | 56 | | 28.87% | 33 | | 21.29% | 58 | | 27.10% | 448 | 16.85% | |
| Moderate (8-19) | | 650 | | 50.90% | 284 | 57.26% | 171 | | 53.11% | 102 | | 52.58% | 82 | | 52.90% | 114 | | 53.27% | 1,403 | 52.78% | |
| Severe (20-35) | | 474 | | 37.12% | 126 | 25.40% | 89 | | 27.64% | 36 | | 18.56% | 40 | | 25.81% | 42 | | 19.63% | 807 | 30.36% | |
| Last pre-treatment IPSS | | 1,533 | | 100.00% | 541 | 100.00% | 350 | | 100.00% | 215 | | 100.00% | 171 | | 100.00% | 239 | | 100.00% | 3,049 | 100.00% | |
| Mean | | 16.47 | | 8.46 | 14.89 | 7.69 | 15.15 | | 7.86 | 13.73 | | 8.21 | 14.94 | | 7.84 | 13.55 | | 7.73 | 15.53 | 8.21 | |
| Median | | 16 | | (9, 23) | 14 | (9, 20) | 14 | | (10, 20) | 13 | | (8, 18) | 14 | | (10, 20) | 14 | | (8, 19) | 15 | (9, 21) | |
| Mild (0-7) | | 182 | | 11.87% | 73 | 13.49% | 51 | | 14.57% | 48 | | 22.33% | 30 | | 17.54% | 48 | | 20.08% | 432 | 14.17% | |
| Moderate (8-19) | | 776 | | 50.62% | 322 | 59.52% | 209 | | 59.71% | 115 | | 53.49% | 96 | | 56.14% | 143 | | 59.83% | 1,661 | 54.48% | |
| Severe (20-35) | | 575 | | 37.51% | 146 | 26.99% | 90 | | 25.71% | 52 | | 24.19% | 45 | | 26.32% | 48 | | 20.08% | 956 | 31.35% | |
| *3 months post-treatment* | | | | | | | | | | | | | | | | | | | | | |
| 3-month follow-up IPSS | | 1,264 | | 82.45% | 456 | 84.29% | 297 | | 84.86% | 168 | | 78.14% | 146 | | 85.38% | 210 | | 87.87% | 2,541 | 83.34% | |
| Mean | | 15.11 | | 8.44 | 13.71 | 7.76 | 14.15 | | 7.36 | 13.04 | | 7.90 | 14.53 | | 7.75 | 12.68 | | 7.49 | 14.38 | 8.08 | |
| Median | | 14 | | (8, 21) | 13 | (8, 19) | 14 | | (9, 18) | 12 | | (7, 17) | 14 | | (10, 19) | 12 | | (8, 17) | 14 | (8, 20) | |
| Mild (0-7) | | 208 | | 16.46% | 77 | 16.89% | 48 | | 16.16% | 42 | | 25.00% | 26 | | 17.81% | 51 | | 24.29% | 452 | 17.79% | |
| Moderate (8-19) | | 664 | | 52.53% | 276 | 60.53% | 188 | | 63.30% | 91 | | 54.17% | 85 | | 58.22% | 126 | | 60.00% | 1,430 | 56.28% | |
| Severe (20-35) | | 392 | | 31.01% | 103 | 22.59% | 61 | | 20.54% | 35 | | 20.83% | 35 | | 23.97% | 33 | | 15.71% | 659 | 25.93% | |
| Change from baseline IPSS | |  | |  |  |  |  | |  |  | |  |  | |  |  | |  |  |  | |
| Mean, SD | | -1.38 | | 5.08 | -1.23 | 4.94 | -1.08 | | 4.79 | -1.17 | | 3.96 | -0.58 | | 2.87 | -1.02 | | 4.25 | -1.23 | 4.78 | |
| Median, IQR | | 0 | | (0, 0) | 0 | (0, 0) | 0 | | (0, 0) | 0 | | (0, 0) | 0 | | (0, 0) | 0 | | (0, 0) | 0 | (0, 0) | |
| Clinic visits per patient | | 1,533 | | 100.00% | 541 | 100.00% | 350 | | 100.00% | 215 | | 100.00% | 171 | | 100.00% | 239 | | 100.00% | 3,049 | 100.00% | |
| Mean, SD | | 2.63 | | 2.15 | 2.40 | 1.78 | 2.22 | | 1.53 | 2.17 | | 1.74 | 2.31 | | 1.42 | 2.31 | | 1.51 | 2.46 | 1.92 | |
| Median, IQR | | 2 | | (1, 3) | 2 | (1, 3) | 2 | | (1, 3) | 2 | | (1, 3) | 2 | | (1, 3) | 2 | | (1, 3) | 2 | (1, 3) | |
| *6 months post-treatment* | | | | | | | | | | | | | | | | | | | | | |
| 6-month follow-up IPSS | | 832 | | 54.27% | 257 | 47.50% | 162 | | 46.29% | 121 | | 56.28% | 84 | | 49.12% | 106 | | 44.35% | 1,562 | 51.23% | |
| Mean | | 13.53 | | 8.63 | 12.72 | 7.70 | 12.52 | | 8.09 | 12.09 | | 7.52 | 12.31 | | 7.69 | 12.25 | | 7.26 | 13.03 | 8.21 | |
| Median | | 13 | | (8, 20) | 12 | (7, 18) | 12 | | (6, 18) | 12 | | (7, 17) | 11 | | (8, 17) | 12 | | (8, 16) | 12 | (7, 18) | |
| Mild (0-7) | | 205 | | 24.64% | 65 | 25.29% | 45 | | 27.78% | 36 | | 29.75% | 20 | | 23.81% | 26 | | 24.53% | 397 | 25.42% | |
| Moderate (8-19) | | 411 | | 49.40% | 141 | 54.86% | 89 | | 54.94% | 64 | | 52.89% | 53 | | 63.10% | 63 | | 59.43% | 821 | 52.56% | |
| Severe (20-35) | | 216 | | 25.96% | 51 | 19.84% | 28 | | 17.28% | 21 | | 17.36% | 11 | | 13.10% | 17 | | 16.04% | 344 | 22.02% | |
| Change from baseline IPSS | |  | |  |  |  |  | |  |  | |  |  | |  |  | |  |  |  | |
| Mean, SD | | -3.13 | | 7.11 | -1.94 | 6.38 | -2.77 | | 6.26 | -1.60 | | 6.32 | -2.63 | | 6.22 | -1.88 | | 6.90 | -2.67 | 6.80 | |
| Median, IQR | | 0 | | (-3, 0) | 0 | (-2, 0) | 0 | | (-1, 0) | 0 | | (0, 0) | 0 | | (-3, 0) | 0 | | (0, 0) | 0 | (-2, 0) | |
| Clinic visits per patient | | 1,533 | | 100.00% | 541 | 100.00% | 350 | | 100.00% | 215 | | 100.00% | 171 | | 100.00% | 239 | | 100.00% | 3,049 | 100.00% | |
| Mean, SD | | 1.43 | | 2.80 | 1.13 | 2.65 | 1.17 | | 3.29 | 1.29 | | 2.23 | 1.30 | | 2.52 | 1.01 | | 2.28 | 1.30 | 2.75 | |
| Median, IQR | | 1 | | (0, 2) | 1 | (0, 1) | 1 | | (0, 1) | 1 | | (0, 1) | 1 | | (0, 1) | 1 | | (0, 1) | 1 | (0, 1) | |
| *1-year post-treatment* | | | | | | | | | | | | | | | | | | | | | |
| 1-year follow-up IPSS | | 768 | | 50.10% | 223 | 41.22% | 151 | | 43.14% | 94 | | 43.72% | 66 | | 38.60% | 83 | | 34.73% | 1,385 | 45.42% | |
| Mean | | 12.40 | | 8.68 | 11.65 | 7.96 | 12.31 | | 7.27 | 10.39 | | 7.32 | 11.80 | | 7.36 | 10.10 | | 7.08 | 11.97 | 8.20 | |
| Median | | 11 | | (5, 19) | 11 | (5, 16) | 12 | | (7, 17) | 10 | | (4, 14) | 11 | | (6, 16) | 11 | | (4, 15) | 11 | (5, 17) | |
| Mild (0-7) | | 237 | | 30.86% | 65 | 29.15% | 39 | | 25.83% | 39 | | 41.49% | 20 | | 30.30% | 30 | | 36.14% | 430 | 31.05% | |
| Moderate (8-19) | | 359 | | 46.74% | 123 | 55.16% | 86 | | 56.95% | 46 | | 48.94% | 37 | | 56.06% | 49 | | 59.04% | 700 | 50.54% | |
| Severe (20-35) | | 172 | | 22.40% | 35 | 15.70% | 26 | | 17.22% | 9 | | 9.57% | 9 | | 13.64% | 4 | | 4.82% | 255 | 18.41% | |
| Change from baseline IPSS | |  | |  |  |  |  | |  |  | |  |  | |  |  | |  |  |  | |
| Mean, SD | | -3.96 | | 7.99 | -3.34 | 6.96 | -2.33 | | 6.64 | -2.85 | | 6.84 | -2.82 | | 6.09 | -1.45 | | 7.61 | -3.40 | 7.54 | |
| Median, IQR | | 0 | | (-6, 0) | 0 | (-7, 0) | 0 | | (-2, 0) | 0 | | (-4, 0) | 0 | | (-3, 0) | 0 | | (0, 0) | 0 | (-5, 0) | |
| Clinic visits per patient | | 1,533 | | 100.00% | 541 | 100.00% | 350 | | 100.00% | 215 | | 100.00% | 171 | | 100.00% | 239 | | 100.00% | 3,049 | 100.00% | |
| Mean, SD | | 1.83 | | 3.38 | 1.31 | 2.10 | 1.17 | | 2.00 | 1.13 | | 1.71 | 1.18 | | 2.01 | 1.41 | | 4.30 | 1.54 | 2.99 | |
| Median, IQR | | 1 | | (0, 2) | 1 | (0, 2) | 1 | | (0, 1) | 1 | | (0, 1) | 1 | | (0, 1) | 0 | | (0, 1) | 1 | (0, 2) | |
| *2-year post-treatment* | | | | | | | | | | | | | | | | | | | | | |
| 2-year follow-up IPSS | | 812 | | 52.97% | 239 | 44.18% | 143 | | 40.86% | 98 | | 45.58% | 74 | | 43.27% | 67 | | 28.03% | 1,433 | 47.00% | |
| Mean | | 11.97 | | 8.82 | 10.45 | 7.75 | 11.85 | | 7.15 | 11.10 | | 7.50 | 12.14 | | 7.23 | 10.69 | | 8.39 | 11.59 | 8.32 | |
| Median | | 11 | | (4, 18) | 10 | (4, 15) | 12 | | (6, 17) | 10 | | (5, 17) | 11 | | (7, 17) | 11 | | (3, 15) | 11 | (4, 17) | |
| Mild (0-7) | | 281 | | 34.61% | 90 | 37.66% | 41 | | 28.67% | 38 | | 38.78% | 23 | | 31.08% | 26 | | 38.81% | 499 | 34.82% | |
| Moderate (8-19) | | 358 | | 44.09% | 120 | 50.21% | 79 | | 55.24% | 47 | | 47.96% | 37 | | 50.00% | 33 | | 49.25% | 674 | 47.03% | |
| Severe (20-35) | | 173 | | 21.31% | 29 | 12.13% | 23 | | 16.08% | 13 | | 13.27% | 14 | | 18.92% | 8 | | 11.94% | 260 | 18.14% | |
| Change from baseline IPSS | |  | |  |  |  |  | |  |  | |  |  | |  |  | |  |  |  | |
| Mean, SD | | -4.76 | | 8.21 | -3.99 | 7.5 | -2.00 | | 6.15 | -2.32 | | 8.39 | -3.68 | | 5.86 | -0.81 | | 8.26 | -3.95 | 7.90 | |
| Median, IQR | | 0 | | (-9, 0) | 0 | (-8, 0) | 0 | | (-3, 0) | 0 | | (-4, 0) | 0 | | (-7, 0. | 0 | | (0, 0) | 0 | (-7, 0) | |
| Clinic visits per patient | | 1,533 | | 100.00% | 541 | 100.00% | 350 | | 100.00% | 215 | | 100.00% | 171 | | 100.00% | 239 | | 100.00% | 3,049 | 100.00% | |
| Mean, SD | | 2.39 | | 3.65 | 1.75 | 3.24 | 1.53 | | 2.73 | 2.08 | | 3.60 | 1.43 | | 2.51 | 1.17 | | 2.39 | 2.01 | 3.37 | |
| Median, IQR | | 1 | | (1, 3) | 1 | (0, 2) | 1 | | (0, 2) | 1 | | (0, 3) | 1 | | (0, 2) | 0 | | (0, 1) | 1 | (0, 2) | |
| *3-year post-treatment* | | | | | | | | | | | | | | | | | | | | | |
| 3-year follow-up IPSS | | 616 | | 40.18% | 188 | 34.75% | 115 | | 32.86% | 65 | | 30.23% | 40 | | 23.39% | 22 | | 9.21% | 1,046 | 34.31% | |
| Mean | | 12.05 | | 8.46 | 10.37 | 7.68 | 11.91 | | 7.57 | 9.97 | | 6.69 | 11.13 | | 7.14 | 8.82 | | 8.00 | 11.50 | 8.10 | |
| Median | | 11 | | (4, 18) | 10 | (3, 15) | 12 | | (6, 17) | 10 | | (4, 14) | 10 | | (6, 16) | 9 | | (1, 13) | 11 | (4, 17) | |
| Mild (0-7) | | 203 | | 32.95% | 70 | 37.23% | 37 | | 32.17% | 24 | | 36.92% | 13 | | 32.50% | 10 | | 45.45% | 357 | 34.13% | |
| Moderate (8-19) | | 283 | | 45.94% | 94 | 50.00% | 60 | | 52.17% | 37 | | 56.92% | 22 | | 55.00% | 11 | | 50.00% | 507 | 48.47% | |
| Severe (20-35) | | 130 | | 21.10% | 24 | 12.77% | 18 | | 15.65% | 4 | | 6.15% | 5 | | 12.50% | 1 | | 4.55% | 182 | 17.40% | |
| Change from baseline IPSS | |  | |  |  |  |  | |  |  | |  |  | |  |  | |  |  |  | |
| Mean, SD | | -4.50 | | 8.59 | -4.26 | 7.90 | -1.55 | | 5.91 | -1.95 | | 6.59 | -4.23 | | 8.05 | -0.41 | | 4.81 | -3.88 | 8.08 | |
| Median, IQR | | 0 | | (-8, 0) | 0 | (-8, 0) | 0 | | (-2, 0) | 0 | | (-3, 0) | 0 | | (-11, 0) | 0 | | (0, 0) | 0 | (-7, 0) | |
| Clinic visits per patient | | 1,533 | | 100.00% | 541 | 100.00% | 350 | | 100.00% | 215 | | 100.00% | 171 | | 100.00% | 239 | | 100.00% | 3,049 | 100.00% | |
| Mean, SD | | 4.31 | | 7.51 | 3.35 | 6.18 | 3.56 | | 7.88 | 2.57 | | 4.76 | 1.88 | | 3.51 | 0.35 | | 1.19 | 3.49 | 6.78 | |
| Median, IQR | | 1 | | (0, 6) | 0 | (0, 5) | 0 | | (0, 4) | 0 | | (0, 3) | 0 | | (0, 2) | 0 | | (0, 0) | 0 | (0, 4) | |

IQR = Interquartile range; SD = standard deviation; IPSS = International Prostate Symptom Score.

Table 1c. Change in IPSS scores for patients pursuing MIST for BPH stratified by time-to-treatment.

|  | | *<12 months from diagnosis*  *N = 1,741* | | | *12-23 months from diagnosis*  *N = 665* | | | *24-35 months from diagnosis*  *N = 501* | | | | *36-47 months from diagnosis*  *N = 418* | | | *48-59 months from diagnosis*  *N = 321* | | | *>60 months from diagnosis*  *N = 373* | | | *All patients*  *N = 4,019* | | |
| --- | --- | --- | --- | --- | --- | --- | --- | --- | --- | --- | --- | --- | --- | --- | --- | --- | --- | --- | --- | --- | --- | --- | --- |
|  | | ***n*** | | ***SD/IQR or %*** | ***n*** | | ***SD/IQR or %*** | ***n*** | | ***SD/IQR or %*** | | ***n*** | | ***SD/IQR or %*** | ***n*** | | ***SD/IQR or %*** | ***n*** | | ***SD/IQR or %*** | ***n*** | ***SD/IQR or %*** | |
| Mean number of IPSS evaluations between diagnosis and treatment | 3.17 | | 1.84 | | | 4.56 | 3.13 | | 5.30 | | 3.36 | | 5.53 | 3.67 | | 6.06 | 4.07 | | 6.91 | 5.18 | 4.49 | | 3.37 |
| Mean time between diagnosis and treatment (months) | 4.46 | | 3.04 | | | 17.43 | 3.42 | | 29.49 | | 3.46 | | 41.78 | 3.39 | | 53.5 | 3.52 | | 74.73 | 11.34 | 24.05 | | 23.22 |
| *Before treatment* | | | | | | | | | | | | | | | | | | | | | | | |
| First pre-treatment IPSS | | 1,430 | | 82.14% | 588 | | 88.42% | 454 | | 90.62% | | 369 | | 88.28% | 283 | | 88.16% | 345 | | 92.49% | 3,469 | 86.32% | |
| Mean | | 18.09 | | 7.47 | 15.24 | | 7.50 | 15.55 | | 7.38 | | 14.55 | | 7.62 | 13.44 | | 7.51 | 13.60 | | 7.61 | 16.07 | 7.70 | |
| Median | | 18 | | (13, 23) | 15 | | (10, 20) | 15 | | (10, 21) | | 14 | | (8, 20) | 12 | | (8, 19) | 13 | | (8, 18) | 16 | (10, 22) | |
| Mild (0-7) | | 107 | | 7.48% | 88 | | 14.97% | 58 | | 12.78% | | 67 | | 18.16% | 66 | | 23.32% | 78 | | 22.61% | 464 | 13.38% | |
| Moderate (8-19) | | 693 | | 48.46% | 324 | | 55.10% | 260 | | 57.27% | | 196 | | 53.12% | 154 | | 54.42% | 191 | | 55.36% | 1,818 | 52.41% | |
| Severe (20-35) | | 630 | | 44.06% | 176 | | 29.93% | 136 | | 29.96% | | 106 | | 28.73% | 63 | | 22.26% | 76 | | 22.03% | 1,187 | 34.22% | |
| Last pre-treatment IPSS | | 1,741 | | 100.00% | 665 | | 100.00% | 501 | | 100.00% | | 418 | | 100.00% | 321 | | 100.00% | 373 | | 100.00% | 4,019 | 100.00% | |
| Mean | | 17.75 | | 7.61 | 15.38 | | 7.39 | 15.63 | | 7.31 | | 15.20 | | 7.68 | 14.77 | | 7.27 | 13.95 | | 7.40 | 16.24 | 7.62 | |
| Median | | 18 | | (13, 23) | 15 | | (10, 20) | 15 | | (11, 21) | | 14 | | (10, 20) | 14 | | (10, 20) | 13 | | (10, 18) | 16 | (11, 21) | |
| Mild (0-7) | | 156 | | 8.96% | 85 | | 12.78% | 54 | | 10.78% | | 57 | | 13.64% | 49 | | 15.26% | 66 | | 17.69% | 467 | 11.62% | |
| Moderate (8-19) | | 854 | | 49.05% | 393 | | 59.10% | 296 | | 59.08% | | 251 | | 60.05% | 187 | | 58.26% | 223 | | 59.79% | 2,204 | 54.84% | |
| Severe (20-35) | | 731 | | 41.99% | 187 | | 28.12% | 151 | | 30.14% | | 110 | | 26.32% | 85 | | 26.48% | 84 | | 22.52% | 1,348 | 33.54% | |
| *3 months post-treatment* | | | | | | | | | | | | | | | | | | | | | | | |
| 3-month follow-up IPSS | | 1,497 | | 85.99% | 577 | | 86.77% | 444 | | 88.62% | | 356 | | 85.17% | 281 | | 87.54% | 322 | | 86.33% | 3,477 | 86.51% | |
| Mean | | 15.50 | | 7.86 | 13.33 | | 7.39 | 13.77 | | 7.58 | | 13.27 | | 7.36 | 13.19 | | 7.03 | 12.54 | | 7.66 | 14.23 | 7.69 | |
| Median | | 15 | | (10, 21) | 13 | | (8, 18) | 13 | | (9, 19) | | 13 | | (9, 18) | 12 | | (8, 18) | 12 | | (7, 17. | 14 | (9, 19) | |
| Mild (0-7) | | 240 | | 16.03% | 127 | | 22.01% | 88 | | 19.82% | | 68 | | 19.10% | 59 | | 20.99% | 83 | | 25.78% | 665 | 19.13% | |
| Moderate (8-19) | | 796 | | 53.17% | 338 | | 58.58% | 255 | | 57.43% | | 217 | | 60.96% | 166 | | 59.07% | 187 | | 58.07% | 1,959 | 56.34% | |
| Severe (20-35) | | 461 | | 30.79% | 112 | | 19.41% | 101 | | 22.75% | | 71 | | 19.94% | 56 | | 19.93% | 52 | | 16.15% | 853 | 24.53% | |
| Change from baseline IPSS | |  | |  |  | |  |  | |  | |  | |  |  | |  |  | |  |  |  | |
| Mean, SD | | -2.44 | | 6.19 | -2.02 | | 5.30 | -1.98 | | 5.51 | | -2.47 | | 5.89 | -1.80 | | 6.06 | -1.63 | | 4.85 | -2.19 | 5.81 | |
| Median, IQR | | 0 | | (-3, 0) | 0 | | (-2, 0) | 0 | | (-2, 0) | | 0 | | (-3, 0) | 0 | | (-1, 0) | 0 | | (0, 0) | 0 | (-2, 0) | |
| Clinic visits per patient | | 1,741 | | 100.00% | 665 | | 100.00% | 501 | | 100.00% | | 418 | | 100.00% | 321 | | 100.00% | 373 | | 100.00% | 4,019 | 100.00% | |
| Mean, SD | | 2.41 | | 2.89 | 2.30 | | 2.55 | 2.34 | | 2.74 | | 2.36 | | 2.96 | 2.10 | | 2.03 | 2.46 | | 3.65 | 2.36 | 2.85 | |
| Median, IQR | | 2 | | (1, 3) | 2 | | (1, 3) | 2 | | (1, 3) | | 2 | | (1, 3) | 2 | | (1, 3) | 2 | | (1, 3) | 2 | (1, 3) | |
| *6 months post-treatment* | | | | | | | | | | | | | | | | | | | | | | | |
| 6-month follow-up IPSS | | 801 | | 46.01% | 297 | | 44.66% | 232 | | 46.31% | | 189 | | 45.22% | 113 | | 35.20% | 164 | | 43.97% | 1,796 | 44.69% | |
| Mean | | 14.97 | | 7.80 | 12.59 | | 7.20 | 12.38 | | 7.20 | | 12.82 | | 7.55 | 13.19 | | 6.96 | 12.09 | | 7.24 | 13.64 | 7.59 | |
| Median | | 15 | | (10, 20) | 12 | | (8, 17) | 12 | | (7, 17) | | 12 | | (8, 17) | 13 | | (8, 18) | 12 | | (7, 17) | 13 | (9, 19) | |
| Mild (0-7) | | 142 | | 17.73% | 74 | | 24.92% | 60 | | 25.86% | | 41 | | 21.69% | 24 | | 21.24% | 44 | | 26.83% | 385 | 21.44% | |
| Moderate (8-19) | | 442 | | 55.18% | 175 | | 58.92% | 131 | | 56.47% | | 112 | | 59.26% | 65 | | 57.52% | 97 | | 59.15% | 1,022 | 56.90% | |
| Severe (20-35) | | 217 | | 27.09% | 48 | | 16.16% | 41 | | 17.67% | | 36 | | 19.05% | 24 | | 21.24% | 23 | | 14.02% | 389 | 21.66% | |
| Change from baseline IPSS | |  | |  |  | |  |  | |  | |  | |  |  | |  |  | |  |  |  | |
| Mean, SD | | -3.21 | | 6.79 | -2.97 | | 6.34 | -2.72 | | 6.36 | | -2.70 | | 6.30 | -1.79 | | 5.47 | -1.54 | | 5.58 | -2.81 | 6.45 | |
| Median, IQR | | 0 | | (-6, 0) | 0 | | (-5, 0) | 0 | | (-5, 0) | | 0 | | (-6, 0) | 0 | | (-1, 0) | 0 | | (-1, 0) | 0 | (-5, 0) | |
| Clinic visits per patient | | 1,741 | | 100.00% | 665 | | 100.00% | 501 | | 100.00% | | 418 | | 100.00% | 321 | | 100.00% | 373 | | 100.00% | 4,019 | 100.00% | |
| Mean, SD | | 1.1396 | | 2.4393 | 1.018 | | 2.1787 | 0.9441 | | 2.3893 | | 0.9737 | | 1.6684 | 0.7882 | | 1.6729 | 1.0027 | | 2.7848 | 1.0371 | 2.3049 | |
| Median, IQR | | 1 | | (0, 1) | 1 | | (0, 1) | 1 | | (0, 1) | | 1 | | (0, 1) | 0 | | (0, 1) | 0 | | (0, 1) | 1 | (0, 1) | |
| *1-year post-treatment* | | | | | | | | | | | | | | | | | | | | | | | |
| 1-year follow-up IPSS | | 937 | | 53.82% | 318 | | 47.82% | 241 | | 48.10% | | 210 | | 50.24% | 147 | | 45.79% | 160 | | 42.90% | 2,013 | 50.09% | |
| Mean | | 14.12 | | 7.88 | 11.94 | | 6.89 | 12.06 | | 7.26 | | 11.84 | | 7.45 | 11.89 | | 6.66 | 11.29 | | 6.85 | 12.90 | 7.53 | |
| Median | | 13 | | (10, 19) | 12 | | (7, 16) | 11 | | (6, 17) | | 11 | | (7, 16) | 12 | | (7, 15) | 11 | | (6, 16) | 12 | (8, 18) | |
| Mild (0-7) | | 191 | | 20.38% | 91 | | 28.62% | 68 | | 28.22% | | 55 | | 26.19% | 40 | | 27.21% | 48 | | 30.00% | 493 | 24.49% | |
| Moderate (8-19) | | 520 | | 55.50% | 186 | | 58.49% | 133 | | 55.19% | | 123 | | 58.57% | 86 | | 58.50% | 96 | | 60.00% | 1,144 | 56.83% | |
| Severe (20-35) | | 226 | | 24.12% | 41 | | 12.89% | 40 | | 16.60% | | 32 | | 15.24% | 21 | | 14.29% | 16 | | 10.00% | 376 | 18.68% | |
| Change from baseline IPSS | |  | |  |  | |  |  | |  | |  | |  |  | |  |  | |  |  |  | |
| Mean, SD | | -3.58 | | 7.34 | -2.67 | | 6.05 | -2.32 | | 5.88 | | -2.86 | | 6.36 | -2.59 | | 6.19 | -1.81 | | 6.05 | -3.00 | 6.72 | |
| Median, IQR | | 0 | | (-7, 0) | 0 | | (-5, 0) | 0 | | (-5, 0) | | 0 | | (-6, 0) | 0 | | (-6, 0) | 0 | | (-3, 0) | 0 | (-6, 0) | |
| Clinic visits per patient | | 1,741 | | 100.00% | 665 | | 100.00% | 501 | | 100.00% | | 418 | | 100.00% | 321 | | 100.00% | 373 | | 100.00% | 4,019 | 100.00% | |
| Mean, SD | | 1.67 | | 2.76 | 1.35 | | 2.35 | 1.17 | | 1.75 | | 1.42 | | 2.01 | 1.06 | | 1.36 | 1.10 | | 1.96 | 1.43 | 2.36 | |
| Median, IQR | | 1 | | (0, 2) | 1 | | (0, 2) | 1 | | (0, 2) | | 1 | | (0, 2) | 1 | | (0, 2) | 1 | | (0, 1) | 1 | (0, 2) | |
| *2-year post-treatment* | | | | | | | | | | | | | | | | | | | | | | | |
| 2-year follow-up IPSS | | 927 | | 53.25% | 273 | | 41.05% | 231 | | 46.11% | | 216 | | 51.67% | 136 | | 42.37% | 131 | | 35.12% | 1,914 | 47.62% | |
| Mean | | 13.03 | | 7.85 | 11.90 | | 6.70 | 11.95 | | 6.91 | | 11.44 | | 7.06 | 10.69 | | 6.15 | 11.12 | | 6.89 | 12.26 | 7.36 | |
| Median | | 13 | | (7, 18) | 12 | | (7, 16) | 11 | | (7, 16) | | 11 | | (6, 15) | 10 | | (6, 14) | 11 | | (6, 15) | 12 | (7, 17) | |
| Mild (0-7) | | 233 | | 25.13% | 76 | | 27.84% | 64 | | 27.71% | | 61 | | 28.24% | 41 | | 30.15% | 40 | | 30.53% | 515 | 26.91% | |
| Moderate (8-19) | | 510 | | 55.02% | 163 | | 59.71% | 132 | | 57.14% | | 129 | | 59.72% | 82 | | 60.29% | 76 | | 58.02% | 1,092 | 57.05% | |
| Severe (20-35) | | 184 | | 19.85% | 34 | | 12.45% | 35 | | 15.15% | | 26 | | 12.04% | 13 | | 9.56% | 15 | | 11.45% | 307 | 16.04% | |
| Change from baseline IPSS | |  | |  |  | |  |  | |  | |  | |  |  | |  |  | |  |  |  | |
| Mean, SD | | -4.40 | | 7.64 | -3.03 | | 6.24 | -2.39 | | 6.24 | | -3.04 | | 6.18 | -3.39 | | 6.76 | -2.25 | | 6.95 | -3.59 | 7.07 | |
| Median, IQR | | 0 | | (-9, 0) | 0 | | (-6, 0) | 0 | | (-5, 0) | | 0 | | (-6, 0) | -1 | | (-7, 0) | 0 | | (-4, 0) | 0 | (-7, 0) | |
| Clinic visits per patient | | 1,741 | | 100.00% | 665 | | 100.00% | 501 | | 100.00% | | 418 | | 100.00% | 321 | | 100.00% | 373 | | 100.00% | 4,019 | 100.00% | |
| Mean, SD | | 2.24 | | 2.84 | 1.85 | | 3.34 | 1.82 | | 3.17 | | 2.09 | | 2.89 | 2.05 | | 2.91 | 1.50 | | 2.62 | 2.02 | 2.97 | |
| Median, IQR | | 1 | | (0, 3) | 1 | | (0, 2) | 1 | | (0, 2) | | 1 | | (0, 3) | 1 | | (0, 3) | 0 | | (0, 2) | 1 | (0, 3) | |
| *3-year post-treatment* | | | | | | | | | | | | | | | | | | | | | | | |
| 3-year follow-up IPSS | | 650 | | 37.33% | 214 | | 32.18% | 166 | | 33.13% | | 140 | | 33.49% | 87 | | 27.10% | 50 | | 13.40% | 1,307 | 32.52% | |
| Mean | | 13.43 | | 8.04 | 11.81 | | 6.51 | 11.35 | | 6.90 | | 11.19 | | 7.13 | 10.53 | | 5.44 | 10.38 | | 6.83 | 12.35 | 7.44 | |
| Median | | 13 | | (8, 19) | 12 | | (7, 17) | 11 | | (6, 15) | | 11 | | (6, 15) | 10 | | (8, 14) | 10 | | (5, 14) | 12 | (7, 17) | |
| Mild (0-7) | | 156 | | 24.00% | 54 | | 25.23% | 46 | | 27.71% | | 37 | | 26.43% | 20 | | 22.99% | 16 | | 32.00% | 329 | 25.17% | |
| Moderate (8-19) | | 352 | | 54.15% | 138 | | 64.49% | 98 | | 59.04% | | 86 | | 61.43% | 60 | | 68.97% | 29 | | 58.00% | 763 | 58.38% | |
| Severe (20-35) | | 142 | | 21.85% | 22 | | 10.28% | 22 | | 13.25% | | 17 | | 12.14% | 7 | | 8.05% | 5 | | 10.00% | 215 | 16.45% | |
| Change from baseline IPSS | |  | |  |  | |  |  | |  | |  | |  |  | |  |  | |  |  |  | |
| Mean, SD | | -4.08 | | 7.88 | -3.20 | | 6.35 | -2.59 | | 6.01 | | -1.99 | | 7.10 | -2.48 | | 6.05 | -2.96 | | 7.60 | -3.38 | 7.25 | |
| Median, IQR | | 0 | | (-8, 0) | 0 | | (-7, 0) | 0 | | (-6, 0. | | 0 | | (-5, 0) | 0 | | (-6, 0) | 0 | | (-3, 0) | 0 | (-7, 0) | |
| Clinic visits per patient | | 1,741 | | 100.00% | 665 | | 100.00% | 501 | | 100.00% | | 418 | | 100.00% | 321 | | 100.00% | 373 | | 100.00% | 4,019 | 100.00% | |
| Mean, SD | | 3.53 | | 5.79 | 3.40 | | 6.43 | 3.71 | | 6.27 | | 3.45 | | 5.79 | 2.77 | | 4.08 | 0.97 | | 2.54 | 3.22 | 5.68 | |
| Median, IQR | | 1 | | (0, 5) | 1 | | (0, 4) | 1 | | (0, 5) | | 1 | | (0, 5) | 1 | | (0, 4) | 0 | | (0, 1) | 1 | (0, 4) | |

IQR = Interquartile range; SD = standard deviation; IPSS = International Prostate Symptom Score.

Table 2a. Comparison of median IPSS scores between TTT groups undergoing medical treatment over follow-up.

| Follow-Up | <12 months | 12–23 months | 24–35 months | 36–47 months | 48–59 months | ≥5 years | *p*-value* |
| --- | --- | --- | --- | --- | --- | --- | --- |
| 3 months | 14 | 10 | 9 | 10 | 10 | 9 | < 0.001 |
| 6 months | 13 | 8 | 8 | 10 | 10 | 8 | < 0.001 |
| 1 year | 11 | 8 | 8 | 9 | 10 | 8 | < 0.001 |
| 2 years | 11 | 8 | 8 | 8 | 10 | 9 | < 0.001 |
| 3 years | 11 | 8 | 8 | 8 | 8 | 10 | < 0.001 |

*Kruskal-Wallis test use to generate overall *p-*value.

Table 2b. Pairwise comparison of median IPSS scores between TTT groups undergoing medical treatment over follow-up.

| Follow-up | <12mo vs 12-23mo | <12mo vs 24-35mo | <12mo vs 36-47mo | <12mo vs 48-59mo | <12mo vs 5+yr | 12-23mo vs 24-35mo | 12-23mo vs 36-47mo | 12-23mo vs 48-59mo | 12-23mo vs 5+yr | 24-35mo vs 36-47mo | 24-35mo vs 48-59mo | 24-35mo vs 5+yr | 36-47mo vs 48-59mo | 36-47mo vs 5+yr | 48-59mo vs 5+yr |
| --- | --- | --- | --- | --- | --- | --- | --- | --- | --- | --- | --- | --- | --- | --- | --- |
| 3 months | **14 vs 10**  ******* | **14 vs 9**  ******* | **14 vs 10**  ******* | **14 vs 10**  ******* | **14 vs 9**  ******* | 10 vs 9  ns | 10 vs 10  ns | 10 vs 10  ns | **10 vs 9**  ****** | 9 vs 10  ns | 9 vs 10  ns | 9 vs 9  ns | 10 vs 10  ns | 10 vs 9  ns | **10 vs 9**  ****** |
| 6 months | **13 vs 8**  ******* | **13 vs 8**  ******* | **13 vs 10**  ******* | **13 vs 10**  ******* | **13 vs 8**  ******* | 8 vs 8  ns | 8 vs  10  ns | **8 vs 10**  ******* | **8 vs 8**  ******* | 8 vs 10  ns | **8 vs 10**  ****** | **8 vs 8**  ****** | 9 vs 10  ns | **9 vs 8**  ******* | **10 vs 8**  ******* |
| 1 year | **11 vs 8**  ******* | **11 vs 8**  ******* | **11 vs 9**  ******* | **11 vs 10**  ****** | **11 vs 8**  ******* | 8 vs 8  ns | 8 vs 9  ns | **8 vs 10**  ******* | 8 vs 8  ns | 8 vs 9  ns | **8 vs 10**  ******* | 8 vs 8  ns | 9 vs 10  ns | **9 vs 8**  ******* | **10 vs 8**  ******* |
| 2 years | **11 vs 8**  ******* | **11 vs 8**  ******* | **11 vs 8**  ******* | **11 vs 8**  ******* | **11 vs 10**  ******* | 8 vs 8  ns | 8 vs 8  ns | 8 vs 8  ns | 8 vs 10  ns | 8 vs 8  ns | 8 vs 8  ns | 8 vs 10  ns | 8 vs 8  ns | 8 vs 10  ns | 8 vs 10  ns |
| 3 years | **11 vs 8**  ******* | **11 vs 8**  ******* | **11 vs 8**  ******* | **11 vs 8**  ******* | 11 vs 10  ns | 8 vs 8  ns | 8 vs 8  ns | 8 vs 8  ns | **8 vs 10**  ******* | 8 vs 8  ns | 8 vs 8  ns | **8 vs 10**  ******* | 8 vs 8  ns | **8 vs 10**  ******* | **8 vs 10**  ******* |

15 pairs; α = 0.0033 based on Bonferroni correction. * *p* < 0.05. ** *p* < 0.01. ****p* < 0.001. *p*-values determined based on Mann-Whitney U-test.

Table 2c. Comparison of median IPSS score over time within each TTT group pursuing medical treatment.

| Time from diagnosis to treatment | 3 months | 6 months | 1 year | 2 years | 3 years | *p*-value* |
| --- | --- | --- | --- | --- | --- | --- |
| <12 months | 14 | 13 | 11 | 11 | 11 | < 0.001 |
| 12–23 months | 10 | 8 | 8 | 8 | 8 | < 0.001 |
| 24–35 months | 9 | 8 | 8 | 8 | 8 | 0.008 |
| 36–47 months | 10 | 10 | 9 | 8 | 8 | < 0.001 |
| 48–59 months | 10 | 10 | 10 | 10 | 8 | 0.012 |
| ≥5 years | 9 | 8 | 8 | 9 | 10 | < 0.001 |

*Kruskal-Wallis test use to generate overall *p-*value.

Table 2d. Pairwise comparison of median IPSS score over time within each TTT group pursuing medical treatment.

| Time from diagnosis to treatment | 3mo vs 6mo | 3mo vs 1yr | 3mo vs 2yr | 3mo vs 3yr | 6mo vs 1yr | 6mo vs 2yr | 6mo vs 3yr | 1yr vs 2yr | 1yr vs 3yr | 2yr vs 3yr |
| --- | --- | --- | --- | --- | --- | --- | --- | --- | --- | --- |
| <12 months | **14 vs 13**  ******* | **14 vs 11**  ******* | **14 vs 11**  ******* | **14 vs 11**  ******* | **13 vs 11**  ******* | **13 vs 11**  ******* | **13 vs 11**  ******* | **11 vs 11**  ****** | 11 vs 11  ns | 11 vs 11  ns |
| 12–23 months | **10 vs 8**  ******* | **10 vs 8**  ******* | **10 vs 8**  ******* | **10 vs 8**  ******* | 8 vs 8  ns | 8 vs 8  ns | 8 vs 8  ns | 8 vs 8  ns | 8 vs 8  ns | 8 vs 8  ns |
| 24–35 months | 9 vs 8  ns | **9 vs 8**  ******* | 9 vs 8  ns | **9 vs 8**  ****** | 8 vs 8  ns | 8 vs 8  ns | 8 vs 8  ns | 8 vs 8  ns | 8 vs 8  ns | 8 vs 8  ns |
| 36–47 months | 10 vs 10  ns | 10 vs 9  ns | **10 vs 8**  ******* | **10 vs 8**  ****** | 10 vs 9  ns | **10 vs 8**  ******* | **10 vs 8**  ****** | 9 vs 8  ns | 9 vs 8  ns | 8 vs 8  ns |
| 48–59 months | 10 vs 10  ns | 10 vs 10  ns | 10 vs 10  ns | **10 vs 8**  ****** | 10 vs 10  ns | 10 vs 10  ns | 10 vs 8  ns | 10 vs 10  ns | 10 vs 8  ns | **10 vs 8**  ****** |
| ≥5 years | 9 vs 8  ns | **9 vs 8**  ******* | 9 vs 9  ns | 9 vs 10  ns | 8 vs 8  ns | 8 vs 9  ns | **8 vs 10**  ****** | **8 vs 9**  ******* | **8 vs 10**  ******* | 9 vs 10  ns |

10 pairs; α = 0.005 based on Bonferroni correction. * *p* < 0.05. ** *p* < 0.01. ****p* < 0.001. *p*-values determined based on Mann-Whitney U-test.

Table 2e. Comparison of median IPSS scores between TTT groups undergoing surgery over follow-up.

| Follow-up | <12 months | 12–23 months | 24–35 months | 36–47 months | 48–59 months | ≥5 years | *p*-value* |
| --- | --- | --- | --- | --- | --- | --- | --- |
| 3 months | 14 | 13 | 14 | 12 | 14 | 12 | 0.013 |
| 6 months | 13 | 12 | 12 | 12 | 11 | 12 | 0.306 |
| 1 year | 11 | 11 | 12 | 10 | 11 | 11 | 0.034 |
| 2 years | 11 | 10 | 12 | 10 | 11 | 11 | 0.150 |
| 3 years | 11 | 10 | 12 | 10 | 10 | 9 | 0.096 |

*Kruskal-Wallis test use to generate overall *p-*value.

Table 2f. Pairwise comparison of median IPSS scores between TTT groups undergoing surgery over follow-up.

| Follow-up | <12mo vs 12-23mo | <12mo vs 24-35mo | <12mo vs 36-47mo | <12mo vs 48-59mo | <12mo vs 5+yr | 12-23mo vs 24-35mo | 12-23mo vs 36-47mo | 12-23mo vs 48-59mo | 12-23mo vs 5+yr | 24-35mo vs 36-47mo | 24-35mo vs 48-59mo | 24-35mo vs 5+yr | 36-47mo vs 48-59mo | 36-47mo vs 5+yr | 48-59mo vs 5+yr |
| --- | --- | --- | --- | --- | --- | --- | --- | --- | --- | --- | --- | --- | --- | --- | --- |
| 3 months | 14 vs 13  ns | 14 vs 14  ns | 14 vs 12  ns | 14 vs  14  ns | 14 vs 12  ns | 13 vs 14  ns | **13 vs 12**  ****** | 13 vs 14  ns | 13 vs 12  ns | 14 vs 12  ns | 14 vs 14  ns | 14 vs 12  ns | 12 vs 14  ns | 12 vs 12  ns | 14 vs 12  Ns |
| 6 months | 13 vs 12  ns | 13 vs 12  ns | 13 vs 12  ns | 13 vs 11  ns | 13 vs 12  ns | 12 vs 12  ns | 12 vs 12  ns | 12 vs 11  ns | 12 vs 12  ns | 12 vs 12  ns | 12 vs 11  ns | 12 vs 12  ns | 12 vs 11  ns | 12 vs 12  ns | 11 vs 12  Ns |
| 1 year | 11 vs 11  ns | 11 vs 12  ns | 11 vs 10  ns | 11 vs 11  ns | 11 vs 11  ns | 11 vs 12  ns | **11 vs 10**  ****** | 11 vs 11  ns | 11 vs 11  ns | **12 vs 10**  ****** | 12 vs 11  ns | **12 vs 11**  ****** | 10 vs 11  ns | 10 vs 11  ns | 11 vs 11  ns |
| 2 years | 11 vs 10  ns | 11 vs 12  ns | 11 vs 10  ns | 11 vs 11  ns | 11 vs 11  ns | 10 vs 12  ns | 10 vs 10  ns | 10 vs 11  ns | 10 vs 11  ns | 12 vs 10  ns | 12 vs 11  ns | 12 vs 11  ns | 10 vs 11  ns | 10 vs 11  ns | 11 vs 11  ns |
| 3 years | 11 vs 10  ns | 11 vs 12  ns | 11 vs 10  ns | 11 vs 10  ns | 11 vs 9  ns | **10 vs 12**  ****** | 10 vs 10  ns | 10 vs 10  ns | 10 vs 9  ns | 12 vs 10  ns | 12 vs 10  ns | 12 vs 9  ns | 10 vs 10  ns | 10 vs 9  ns | 10 vs 9  ns |

15 pairs; α = 0.0033 based on Bonferroni correction. * *p* < 0.05. ** *p* < 0.01. ****p* < 0.001. *p*-values determined based on Mann-Whitney U-test.

Table 2g. Comparison of median IPSS score over time within each TTT group pursuing surgery.

| Time from diagnosis to treatment | 3 months | 6 months | 1 year | 2 years | 3 years | *p-*value* |
| --- | --- | --- | --- | --- | --- | --- |
| <12 months | 14 | 13 | 11 | 11 | 11 | < 0.001 |
| 12–23 months | 13 | 12 | 11 | 10 | 10 | < 0.001 |
| 24–35 months | 14 | 12 | 12 | 12 | 12 | 0.009 |
| 36–47 months | 12 | 12 | 10 | 10 | 10 | 0.001 |
| 48–59 months | 14 | 11 | 11 | 11 | 10 | < 0.001 |
| ≥5 years | 12 | 12 | 11 | 11 | 9 | 0.059 |

*Kruskal-Wallis test use to generate overall *p-*value.

Table 2h. Pairwise comparison of median IPSS score over time within each within each TTT group pursuing surgery.

| Time from diagnosis to treatment | 3mo vs 6mo | 3mo vs 1yr | 3mo vs 2yr | 3mo vs 3yr | 6mo vs 1yr | 6mo vs 2yr | 6mo vs 3yr | 1yr vs 2yr | 1yr vs 3yr | 2yr vs 3yr |
| --- | --- | --- | --- | --- | --- | --- | --- | --- | --- | --- |
| <12 months | **14 vs 13**  ****** | **14 vs 11**  ******* | **14 vs 11**  ******* | **14 vs 11**  ******* | **13 vs 11**  ******* | **13 vs 11**  ******* | **13 vs 11**  ******* | 11 vs 11  ns | 11 vs 11  ns | 11 vs 11  ns |
| 12–23 months | 13 vs 12  ns | **13 vs 11**  ******* | **13 vs 10**  ******* | 13 vs 10  ns | **12 vs 11**  ****** | **12 vs 10**  ******* | 12 vs 10  ns | 11 vs 10  ns | 11 vs 10  ns | 10 vs 10  ns |
| 24–35 months | **14 vs 12**  ****** | 14 vs 12  ns | 14 vs 12  ns | **14 vs 12**  ****** | 12 vs 12  ns | 12 vs 12  ns | 12 vs 12  ns | 12 vs 12  ns | 12 vs 12  ns | 12 vs 12  ns |
| 36–47 months | 12 vs 12  ns | **12 vs 10**  ******* | 12 vs 10  ns | **12 vs 10**  ****** | **12 vs 10**  ****** | 12 vs 10  ns | 12 vs 10  ns | 10 vs 10  ns | 10 vs 10  ns | 10 vs 10  ns |
| 48–59 months | **14 vs 11**  ****** | **14 vs 11**  ****** | **14 vs 11**  ****** | **14 vs 10**  ****** | 11 vs 11  ns | 11 vs 11  ns | 11 vs 10  ns | 11 vs 11  ns | 11 vs 10  ns | 11 vs 10  ns |
| ≥5 years | 12 vs 12  ns | 12 vs 11  ns | 12 vs 11  ns | **12 vs 9**  ****** | 12 vs 11  ns | 12 vs 11  ns | **12 vs 9**  ****** | 11 vs 11  ns | 11 vs 9  ns | 11 vs 9  ns |

10 pairs; α = 0.005 based on Bonferroni correction. * *p* < 0.05. ** *p* < 0.01. ****p* < 0.001.

Table 2i. Comparison of median IPSS scores between TTT groups undergoing MIST over follow-up.

| Follow-up | <12 months | 12–23 months | 24–35 months | 36–47 months | 48–59 months | ≥5 years | *p*-value* |
| --- | --- | --- | --- | --- | --- | --- | --- |
| 3 months | 15 | 13 | 13 | 13 | 12 | 12 | < 0.001 |
| 6 months | 15 | 12 | 12 | 12 | 13 | 12 | < 0.001 |
| 1 year | 13 | 12 | 11 | 11 | 12 | 11 | < 0.001 |
| 2 years | 13 | 12 | 11 | 11 | 10 | 11 | < 0.001 |
| 3 years | 13 | 12 | 11 | 11 | 10 | 10 | < 0.001 |

*Kruskal-Wallis test use to generate overall *p-*value.

Table. 2j. Pairwise comparison of median IPSS scores between TTT groups undergoing MIST over follow-up.

| Follow-up | <12mo vs 12-23mo | <12mo vs 24-35mo | <12mo vs 36-47mo | <12mo vs 48-59mo | <12mo vs 5+yr | 12-23mo vs 24-35mo | 12-23mo vs 36-47mo | 12-23mo vs 48-59mo | 12-23mo vs 5+yr | 24-35mo vs 36-47mo | 24-35mo vs 48-59mo | 24-35mo vs 5+yr | 36-47mo vs 48-59mo | 36-47mo vs 5+yr | 48-59mo vs 5+yr |
| --- | --- | --- | --- | --- | --- | --- | --- | --- | --- | --- | --- | --- | --- | --- | --- |
| 3 months | **15 vs 13**  ******* | **15 vs 13**  ******* | **15 vs 13**  ******* | **15 vs 12**  ******* | **15 vs 12**  ******* | 13 vs 13  ns | 13 vs 13  ns | **13 vs 12**  ****** | 13 vs 12  ns | 13 vs 13  ns | **13 vs 12**  ******* | 13 vs 12  ns | 13 vs 12  ns | 13 vs 12  ns | 12 vs 12  ns |
| 6 months | **15 vs 12**  ******* | **15 vs 12**  ******* | **15 vs 12**  ******* | **15 vs 13**  ******* | **15 vs 12**  ******* | 12 vs 12  ns | 12 vs 12  ns | 12 vs 13  ns | 12 vs 12  ns | 12 vs 12  ns | 12 vs 13  ns | 12 vs 12  ns | 12 vs 13  ns | 12 vs 12  ns | 13 vs 12  ns |
| 1 year | **13 vs 12**  ****** | **13 vs 11**  ******* | 13 vs 11  ns | 13 vs 12  ns | **13 vs 11**  ****** | 12 vs 11  ns | 12 vs 11  ns | 12 vs 12  ns | 12 vs 11  ns | 11 vs 11  ns | 11 vs 12  ns | 11 vs 11  ns | 11 vs 12  ns | 11 vs 11  ns | 12 vs 11  ns |
| 2 years | **13 vs 12**  ******* | **13 vs 11**  ******* | **13 vs 11**  ******* | **13 vs 10**  ******* | 13 vs 11  ns | 12 vs 11  ns | 12 vs 11  ns | 12 vs 10  ns | 12 vs 11  ns | 11 vs 11  ns | 11 vs 10  ns | 11 vs 11  ns | 11 vs 10  ns | 11 vs 11  ns | 10 vs 11  ns |
| 3 years | 13 vs 12  ns | **13 vs 11**  ****** | 13 vs11  ns | **13 vs 10**  ******* | **13 vs 10**  ******* | 12 vs 11  ns | 12 vs 11  ns | **12 vs 10**  ******* | **12 vs 10**  ****** | 11 vs 11  ns | 11 vs 10  ns | 11 vs 10  ns | 11 vs 10  ns | 11 vs 10  ns | 10 vs 10  ns |

15 pairs; α = 0.0033 based on Bonferroni correction. * *p* < 0.05. ** *p* < 0.01. ****p* < 0.001.

Table 2k. Comparison of median IPSS score over time within each TTT group pursuing MIST.

| Time from diagnosis to treatment | 3 months | 6 months | 1 year | 2 years | 3 years | *p*-value* |
| --- | --- | --- | --- | --- | --- | --- |
| <12 months | 15 | 15 | 13 | 13 | 13 | < 0.001 |
| 12–23 months | 13 | 12 | 12 | 12 | 12 | < 0.001 |
| 24–35 months | 13 | 12 | 11 | 11 | 11 | < 0.001 |
| 36–47 months | 13 | 12 | 11 | 11 | 11 | 0.003 |
| 48–59 months | 12 | 13 | 12 | 10 | 10 | < 0.001 |
| ≥5 years | 12 | 12 | 11 | 11 | 10 | 0.312 |

*Kruskal-Wallis test use to generate overall *p-*value.

Table 2l. Pairwise comparison of median IPSS score over time within each TTT group pursuing MIST.

| TTT group | 3mo vs 6mo | 3mo vs 1yr | 3mo vs 2yr | 3mo vs 3yr | 6mo vs 1yr | 6mo vs 2yr | 6mo vs 3yr | 1yr vs 2yr | 1yr vs 3yr | 2yr vs 3yr |
| --- | --- | --- | --- | --- | --- | --- | --- | --- | --- | --- |
| <12 months | 15 vs 15  ns | **15 vs 13**  ******* | **15 vs 13**  ******* | **15 vs 13**  ******* | **15 vs 13**  ******* | **15 vs 13**  ******* | **15 vs 13**  ******* | 13 vs 13  ns | 13 vs 13  ns | 13 vs 13  ns |
| 12–23 months | **13 vs 12**  ****** | **13 vs 12**  ****** | **13 vs 12**  ******* | 13 vs 12  ns | 12 vs 12  ns | 12 vs 12  ns | 12 vs 12  ns | 12 vs 12  ns | 12 vs 12  ns | 12 vs 12  ns |
| 24–35 months | 13 vs 12  ns | **13 vs 11**  ******* | **13 vs 11**  ****** | 13 vs 11  ns | **12 vs 11**  ****** | 12 vs 11  ns | 12 vs 11  ns | 11 vs 11  ns | 11 vs 11  ns | 11 vs 11  ns |
| 36–47 months | 13 vs 12  ns | 13 vs 11  ns | **13 vs 11**  ******* | **13 vs 11**  ****** | 12 vs 11  ns | 12 vs 11  ns | 12 vs 11  ns | 11 vs 11  ns | 11 vs 11  ns | 11 vs 11  ns |
| 48–59 months | 12 vs 13  ns | 12 vs 12  ns | 12 vs 10  ns | **12 vs 10**  ****** | 13 vs 12  ns | 13 vs 10  ns | **13 vs 10**  ****** | **12 vs 10**  ****** | **12 vs 10**  ******* | 10 vs 10  ns |
| ≥5 years | 12 vs 12  ns | 12 vs 11  ns | 12 vs 11  ns | 12 vs 10  ns | 12 vs 11  ns | 12 vs 11  ns | 12 vs 10  ns | 11 vs 11  ns | 11 vs 10  ns | 11 vs 10  ns |

10 pairs; α = 0.005 based on Bonferroni correction. * *p* < 0.05. ** *p* < 0.01. ****p* < 0.001.

Table 3a. Percentage change in mild LUTS after 3 months from initiating medical treatment for BPH stratified by TTT.

| **Time-to-treatment** | **Mild IPSS Frequency (%)** | **Moderate + Severe Frequency (%)** | **Δ Mild** | ***p*-value** |
| --- | --- | --- | --- | --- |
| **<12mo** |  |  | -0.8 | 0.281 |
| **Baseline (*n* = 7,726)** | 18.5 | 81.5 |  |  |
| **3-month follow-up (*n* = 4,120)** | 17.7 | 82.3 |  |  |
| **12–23mo** |  |  | -2.7 | 0.182 |
| **Baseline (*n* = 1,149)** | 23.7 | 76.3 |  |  |
| **3-month follow-up (*n* = 606)** | 21.0 | 79.0 |  |  |
| **24–35mo** |  |  | -0.5 | 0.858 |
| **Baseline (*n* = 946)** | 25.6 | 74.4 |  |  |
| **3-month follow-up (*n* = 414)** | 25.1 | 74.9 |  |  |
| **36–47mo** |  |  | -6.0 | 0.025 |
| **Baseline (*n* = 747)** | 23.0 | 77.0 |  |  |
| **3-month follow-up (*n* = 335)** | 17.0 | 83.0 |  |  |
| **48–59mo** |  |  | -2.9 | 0.328 |
| **Baseline (*n* = 590)** | 22.0 | 78.0 |  |  |
| **3-month follow-up (*n* = 287)** | 19.2 | 80.8 |  |  |
| **5+yr** |  |  | -3.2 | 0.137 |
| **Baseline (*n* = 1,116)** | 23.3 | 76.7 |  |  |
| **3-month follow-up (*n* = 581)** | 20.1 | 79.9 |  |  |
| **All** |  |  | **-1.7** | 0.005 |
| **Baseline (*n* = 12,574)** | 20.5 | 79.5 |  |  |
| **3-month follow-up (*n* = 6,343)** | 18.8 | 81.2 |  |  |

Table 3b. Percentage change in mild LUTS after 3 months from initiating surgical treatment for BPH stratified by TTT.

| **Time-to-treatment** | **Mild IPSS Frequency (%)** | **Moderate + Severe Frequency (%)** | **Δ Mild** | ***p*-value** |
| --- | --- | --- | --- | --- |
| **<12mo** |  |  | 4.6 | < 0.001 |
| **Baseline (*n* = 1,533)** | 11.9 | 88.1 |  |  |
| **3-month follow-up (*n* = 1,264)** | 16.5 |  |  |  |
| **12–23mo** |  |  | 3.4 | 0.136 |
| **Baseline (*n* = 541)** | 13.5 | 86.5 |  |  |
| **3-month follow-up (*n* = 456)** | 16.9 |  |  |  |
| **24–35mo** |  |  | 1.6 | 0.576 |
| **Baseline (*n* = 350)** | 14.6 | 85.4 |  |  |
| **3-month follow-up (*n* = 297)** | 16.2 |  |  |  |
| **36–47mo** |  |  | 2.7 | 0.540 |
| **Baseline (*n* = 215)** | 22.3 | 77.7 |  |  |
| **3-month follow-up (*n* = 168)** | 25.0 |  |  |  |
| **48–59mo** |  |  | 0.3 | 0.951 |
| **Baseline (*n* = 171)** | 17.5 | 82.5 |  |  |
| **3-month follow-up (*n* = 146)** | 17.8 |  |  |  |
| **5+yr** |  |  | 4.2 | 0.284 |
| **Baseline (*n* = 239)** | 20.1 | 79.9 |  |  |
| **3-month follow-up (*n* = 210)** | 24.3 |  |  |  |
| **All** |  |  | **3.6** | < 0.001 |
| **Baseline (*n* = 3,049)** | 14.2 | 85.8 |  |  |
| **3-month follow-up (*n* = 2,541)** | 17.8 |  |  |  |

Table 3c. Percentage change in mild LUTS after 3 months from initiating MIST for BPH stratified by TTT.

| **Time-to-treatment** | **Mild IPSS Frequency (%)** | **Moderate + Severe Frequency (%)** | **Δ Mild** | ***p*-value** |
| --- | --- | --- | --- | --- |
| **<12mo** |  |  | 7.1 | < 0.001 |
| **Baseline (*n* = 1,741)** | 9.0 | 91.0 |  |  |
| **3-month follow-up (*n* = 1,497)** | 16.0 |  |  |  |
| **12–23mo** |  |  | 9.2 | < 0.001 |
| **Baseline (*n* = 665)** | 12.8 | 87.2 |  |  |
| **3-month follow-up (*n* = 577)** | 22.0 |  |  |  |
| **24–35mo** |  |  | 9.0 | < 0.001 |
| **Baseline (*n* = 501)** | 10.8 | 89.2 |  |  |
| **3-month follow-up (*n* = 444)** | 19.8 |  |  |  |
| **36–47mo** |  |  | 5.5 | 0.0390 |
| **Baseline (*n* = 418)** | 13.6 | 86.4 |  |  |
| **3-month follow-up (*n* = 356)** | 19.1 |  |  |  |
| **48–59mo** |  |  | 5.7 | 0.067 |
| **Baseline (*n* = 321)** | 15.3 | 84.7 |  |  |
| **3-month follow-up (*n* = 281)** | 21.0 |  |  |  |
| **5+yr** |  |  | 8.1 | 0.010 |
| **Baseline (*n* = 373)** | 17.7 | 82.3 |  |  |
| **3-month follow-up (*n* = 322)** | 25.8 |  |  |  |
| **All** |  |  | **7.5** | < 0.001 |
| **Baseline (*n* = 4,019)** | 11.6 | 88.4 |  |  |
| **3-month follow-up (*n* = 3,477)** | 19.1 |  |  |  |

Table 4a. Time-to-event stratified by time from diagnosis to medical treatment for BPH.

|  | <12 months from diagnosis  *N* = 1,549 | | 1 – 3 years from diagnosis  *N* = 273 | >3 years from diagnosis  *N* = 208 | | All  *N* = 2,030 | |  |
| --- | --- | --- | --- | --- | --- | --- | --- | --- |
| Time-to-event (Days) | | Mean ± SD | Mean ± SD | | Mean ± SD | | Mean ± SD | *p*-value |
| Total events | | **525.03 ± 631.84** | **436.67 ± 522.95** | | **317.7 ± 414.62** | | **491.9 ± 602.61** | **<0.001** |
| Bladder irrigation | | 507.29 ± 610.81 | 432.04 ± 500.09 | | 301.1 ± 388.61 | | 475.64 ± 580.63 | <0.001 |
| Catheterization | | 605.33 ± 690.39 | 477.29 ±  554.56 | | 311.82 ± 432.75 | | 556.87 ± 657.21 | <0.001 |

Table 4b. Time-to-event stratified by time from diagnosis to surgical treatment for BPH.

|  | <12 months from diagnosis  *N* = 386 | 1 – 3 years from diagnosis  *N* = 199 | >3 years from diagnosis  *N* = 159 | All  *N* = 744 | |  |
| --- | --- | --- | --- | --- | --- | --- |
| Time-to-event (Days) | Mean ± SD | Mean ± SD | Mean ± SD | | Mean ± SD | *p*-value |
| Total events | **250.01±**  **570.47** | **115.15 ±**  **326.35** | **76.6 ± 240.89** | | **176.88 ±**  **464.01** | **<0.001** |
| Bladder irrigation | 162.98 ± 445.34 | 95.74 ± 332.11 | 29.42 ± 102.84 | | 114.04 ± 365.93 | 0.0022 |
| Catheterization | 437.01 ± 713.69 | 240.67 ±  426.43 | 189.91 ± 360.42 | | 338.69 ± 601.75 | 0.011 |
| Bleeding  requiring  Fulguration | 616.26 ± 772.83 | 731.83 ± 678.67 | 557.33 ± 531.01 | | 634.71±  712.29 | * |
| TURP | 196.85  ± 486.5 | 91.22  ± 273.36 | 54.3  ± 127.58 | | 140.80  ± 392.18 | 0.031 |
| PVP Laser | 397.33  ± 728.2 | 179.35  ± 424.39 | 138.9  ± 389.32 | | 282.22  ± 604.05 | 0.582 |
| Laser Enucleation | 17.29  ± 25.14 | 15.33  ± 29.3 | 3.33  ± 1.97 | | 10.12  ± 19.58 | 0.514 |
| Aquablation | 33.93  ± 90.15 | 5.57  ± 4.2 | 55.75  ± 142.37 | | 33.1  ± 95.82 | 0.131 |

*Small cohort size and underpowered for comparison.

Table 4c. Time-to-event stratified by time from diagnosis to MIST for BPH.

|  | <12 months from diagnosis  *N* = 381 | 1 – 3 years from diagnosis  *N* = 232 | >3 years from diagnosis  *N* = 198 | All  *N* = 811 | |  |
| --- | --- | --- | --- | --- | --- | --- |
| Time-to-event (Days) | Mean ± SD | Mean ± SD | Mean ± SD | | Mean ± SD | *p*-value |
| Total events | **233.82 1±**  **474.54** | **230.91± 502.4** | **147.84± 357.77** | | **212.00±**  **458.36** | **0.079** |
| Bladder irrigation | 216.59± 449.27 | 171.25± 433.68 | 109.78± 298.72 | | 178.23 ±415.15 | 0.043 |
| Catheterization | 321.90± 522.29 | 389.36±  611.94 | 212.96± 403.73 | | 312.90± 525.08 | 0.070 |
| Bleeding  requiring  Fulguration | 814.25± 684.44 | 271.33± 270.76 | 289.00± 489.30 | | 493.80±549.33 | * |
| UroLift | 281.91  ± 520.18 | 259.01  ± 506.51 | 212.37  ± 418.99 | | 261.06  ± 496.91 | 0.036 |
| Rezum | 71.31  ± 196.26 | 163.12  ± 489.38 | 50.63  ± 204.4 | | 91.01  ± 315.09 | 0.048 |

*Small cohort size and underpowered for comparison.

Table 4d. Pairwise comparisons of mean time-to-event stratified by time from diagnosis to treatment.

| Treatment | <12mo vs 1–3yr Sig. | Difference (days) | <12mo vs >3yr Sig. | Difference (days) | 1–3yr vs >3yr Sig. | Difference (days) |
| --- | --- | --- | --- | --- | --- | --- |
| Medical Treatment | ***** *(p=0.013)* | +88 | ******* *(p=<0.001)* | +207 | ****** *(p=0.006)* | +119 |
| Total Surgical Treatment | ns *(p=0.026)* | +115 | ******* *(p=<0.001)* | +232 | ns *(p=0.040)* | +117 |
| TURP | ns *(p=0.075)* | +128 | ******* *(p=<0.001)* | +254 | ns *(p=0.154)* | +127 |
| PVP Laser | ns *(p=0.803)* | +24 | ns *(p=0.026)* | +144 | ns *(p=0.221)* | +119 |
| Laser Enucleation | ns *(p=0.114)* | +264 | ns *(p=0.261)* | +206 | ns *(p=0.666)* | −59 |
| Aquablation | ns *(p=0.845)* | −44 | ****** *(p=0.0016)* | +370 | ns *(p=0.085)* | +415 |
| Total MIST | ns *(p=0.687)* | +21 | ******* *(p=<0.001)* | +172 | ***** *(p=0.011)* | +151 |
| UroLift | ns *(p=0.261)* | +55 | ******* *(p=<0.001)* | +141 | ns *(p=0.140)* | +86 |
| Rezum | ns *(p=0.783)* | −62 | ******* *(p=<0.001)* | +527 | ***** *(p=0.013)* | +589 |

Table 5a. Time to secondary retreatment after index medical treatment for BPH stratified by time from diagnosis to treatment.

|  | | <12 months | | 1–3 years | | | | | >3 years | |
| --- | --- | --- | --- | --- | --- | --- | --- | --- | --- | --- |
| Retreatment | ***n*** | | **Mean ± SD (months)** | | ***n*** | **Mean ± SD (months)** | ***n*** | **Mean ± SD (months)** | | **p-value** |
| OAB Medication | 1,369 | | 22.1 ± 22.6 | | 291 | 20.6 ± 20.4 | 221 | 12.6 ± 12.7 | | <0.001 |
| Surgery — Total | 656 | | 17.0 ± 20.6 | | 112 | 13.2 ± 15.7 | 69 | 9.3 ± 9.3 | | 0.0024 |
| Surgery — TURP | 448 | | 18.3 ± 20.9 | | 65 | 14.2 ± 17.0 | 32 | 10.0 ± 11.2 | | 0.031 |
| Surgery — PVP Laser | 160 | | 12.2 ± 18.5 | | 35 | 11.4 ± 16.6 | 17 | 7.5 ± 6.0 | | 0.582 |
| Surgery — Laser Enucleation | 27 | | 18.7 ± 24.5 | | 9 | 10.0 ± 7.6 | 8 | 11.9 ± 10.0 | | 0.514 |
| Surgery — Aquablation | 45 | | 20.7 ± 19.7 | | 8 | 22.2 ± 18.8 | 12 | 8.6 ± 7.5 | | 0.131 |
| MIST — Total | 912 | | 15.5 ± 18.0 | | 114 | 14.8 ± 17.0 | 100 | 9.8 ± 10.9 | | 0.0088 |
| MIST — UroLift | 850 | | 14.9 ± 17.4 | | 99 | 13.1 ± 14.7 | 88 | 10.3 ± 11.4 | | 0.036 |
| MIST — Rezum | 64 | | 23.8 ± 23.0 | | 15 | 25.9 ± 25.9 | 12 | 6.5 ± 6.2 | | 0.048 |

Table 5b. Time to surgical retreatment after index surgical treatment for BPH stratified by time from diagnosis to treatment.

|  | <12 months | | 1–3 years | | >3 years | | *p*-value |
| --- | --- | --- | --- | --- | --- | --- | --- |
|  | ***n*** | **Mean ±**  **SD (months)** | ***n*** | **Mean ±**  **SD (months)** | ***n*** | **Mean ±**  **SD (months)** |  |
| Total | 47 | 27.3 ± 19.6 | 25 | 27.8 ± 21.5 | 14 | 17.3 ± 15.5 | 0.237 |
| TURP | 34 | 28.9 ± 21.6 | 19 | 29.9 ± 19.7 | 13 | 17.4 ± 16.1 | 0.201 |
| PVP Laser | 11 | 27.0 ± 19.1 | 6 | 21.1 ± 27.1 | 0 | — | 0.645 |

Table 5c. Time to surgical retreatment after index MIST for BPH stratified by time from diagnosis to treatment.

|  | <12 months | | 1–3 years | | >3 years | | *p*-value |
| --- | --- | --- | --- | --- | --- | --- | --- |
|  | **n** | **Mean ±**  **SD (months)** | **n** | **Mean ±**  **SD (months)** | **n** | **Mean ±**  **SD (months)** |  |
| Total | 15 | 42.4 ± 17.3 | 7 | 40.8 ±2 5.2 | 4 | 16.7 ± 14.3 | 0.142 |
| UroLift | 12 | 42.0 ± 17.7 | 6 | 47.0 ± 21.0 | 4 | 16.7 ± 14.3 | 0.104 |

*Cohort size for Rezum too small for meaningful statistical comparison.

Table 5d. Pairwise comparisons of time to secondary treatment after index medical treatment for BPH by time from diagnosis to treatment.

| Comparison by Time-to-Treatment | Difference (months) | p-value | | |
| --- | --- | --- | --- | --- |
| OAB Medications |  |  | | |
| *<12mo vs 1–3yr* | +1.6 | 0.595 | | |
| *<12mo vs >3yr* | +9.6 | <0.001 | | |
| *1–3yr vs >3yr* | +8.0 | <0.001 | | |
| Surgery – Total |  | | |  |
| *<12mo vs 1–3yr* | +3.8 | | | 0.026 |
| *<12mo vs >3yr* | +7.6 | | | <0.001 |
| *1–3yr vs >3yr* | +3.8 | | | 0.040 |
| Surgery – TURP |  | | |  |
| *<12mo vs 1–3yr* | +4.2 | | | 0.075 |
| *<12mo vs >3yr* | +8.3 | | | <0.001 |
| *1–3yr vs >3yr* | +4.2 | | | 0.154 |
| Surgery – PVP Laser |  | | |  |
| *<12mo vs 1–3yr* | +0.8 | | | 0.803 |
| *<12mo vs >3yr* | +4.7 | | | 0.026 |
| *1–3yr vs >3yr* | +3.9 | | | 0.221 |
| Surgery – Laser Enucleation |  | | |  |
| *<12mo vs 1–3yr* | +8.7 | | | 0.114 |
| *<12mo vs >3yr* | +6.8 | | | 0.261 |
| *1–3yr vs >3yr* | −1.9 | | | 0.666 |
| Surgery - Aquablation |  | | |  |
| *<12mo vs 1–3yr* | −1.5 | | | 0.845 |
| *<12mo vs >3yr* | +12.2 | | | 0.0016 |
| *1–3yr vs >3yr* | +13.6 | | | 0.085 |
| MIST – Total |  | | |  |
| *<12mo vs 1–3yr* | +0.7 | | | 0.687 |
| *<12mo vs >3yr* | +5.7 | | | <0.001 |
| *1–3yr vs >3yr* | +5.0 | | | 0.011 |
| MIST - UroLift |  | | |  |
| *<12mo vs 1–3yr* | +1.8 | | | 0.261 |
| *<12mo vs >3yr* | +4.6 | | | <0.001 |
| *1–3yr vs >3yr* | +2.8 | | | 0.140 |
| MIST – Rezum |  | | |  |
| *<12mo vs 1–3yr* | −2.0 | | 0.783 | |
| *<12mo vs >3yr* | +17.3 | | <0.001 | |
| *1–3yr vs >3yr* | +19.3 | | 0.013 | |

Bonferroni correction α = 0.0167.

Table 5e. Pairwise comparisons of time to surgical retreatment after index surgical treatment for BPH by time from diagnosis to treatment.

| Comparison by Time-to-Treatment | Difference (months) | *p*-value |
| --- | --- | --- |
| Total | | |
| *<12mo vs 1–3yr* | −0.5 | 0.919 |
| *<12mo vs >3yr* | +9.9 | 0.058 |
| *1–3yr vs >3yr* | +10.5 | 0.088 |
| TURP | | |
| *<12mo vs 1–3yr* | −1.1 | 0.856 |
| *<12mo vs >3yr* | +11.4 | 0.058 |
| *1–3yr vs >3yr* | +12.5 | 0.059 |
| PVP Laser | | |
| *<12mo vs 1–3yr* | +6.0 | 0.645 |
| *<12mo vs >3yr* | — | — |
| *1–3yr vs >3yr* | — | — |

Bonferroni correction α = 0.0167.

Table 5f. Pairwise comparisons of time to surgical retreatment after index MIST for BPH by time from diagnosis to treatment.

| Comparison by Time-to-Treatment | | Difference (months) | | *p*-value |
| --- | --- | --- | --- | --- |
| Total |  | |  | |
| *<12mo vs 1–3yr* | | +1.6 | | 0.885 |
| *<12mo vs >3yr* | | +25.6 | | 0.025 |
| *1–3yr vs >3yr* | | +24.1 | | 0.074 |
| Urolift | | −5.0 | | 0.629 |
| *<12mo vs 1–3yr* | | +25.2 | | 0.026 |
| *<12mo vs >3yr* | | +30.2 | | 0.027 |
| *1–3yr vs >3yr* | |  | |  |

Bonferroni correction α = 0.0167.

Table 6. Clinicodemographic variables for patients with and without available IPSS scores within AQUA Registry.

|  | | With IPSS  *N* = 131,062 | | | Without IPSS  *N* = 773,549 | | | | | | *p*-value | | |
| --- | --- | --- | --- | --- | --- | --- | --- | --- | --- | --- | --- | --- | --- |
|  | | ***N*** | | **%** | ***N*** | | | | **%** | |  | | |
| Type of Treatment |  | |  | | |  | |  | | | | <0.001 | |
| Any Surgery | | 6,527 | | 4.98% | 21,968 | | | | 2.84% | |  | | |
| No Surgery | | 124,535 | | 95.02% | 751,581 | | | | 97.16% | |  | | |
| Any MIST | | 7,515 | | 5.73% | 12,730 | | | | 1.65% | | <0.001 | | |
| No MIST | | 123,547 | | 94.27% | 760,819 | | | | 98.35% | |  | | |
| Medication | | 53,496 | | 40.82% | 423,148 | | | | 54.70% | | <0.001 | | |
| TURP | | 3,687 | | 2.81% | 14,502 | | | | 1.87% | |  | | |
| PVP | | 2,428 | | 1.85% | 6,755 | | | | 0.87% | |  | | |
| Laser | | 265 | | 0.20% | 510 | | | | 0.07% | |  | | |
| Aquablation | | 147 | | 0.11% | 201 | | | | 0.03% | |  | | |
| UroLift | | 6,072 | | 4.63% | 9,834 | | | | 1.27% | |  | | |
| Rezum | | 1,443 | | 1.10% | 2,896 | | | | 0.37% | |  | | |
| Patient Age at Baseline | | | | | | |  | | |  | | | <0.001 |
| ≥40, <50 | | 4,595 | | 3.51% | 30,881 | | | | 3.99% | |  | | |
| ≥50, <60 | | 23,272 | | 17.76% | 130,635 | | | | 16.89% | |  | | |
| ≥60, <70 | | 50,960 | | 38.88% | 279,801 | | | | 36.17% | |  | | |
| ≥70, <80 | | 40,747 | | 31.09% | 242,527 | | | | 31.35% | |  | | |
| ≥80 | | 11,488 | | 8.77% | 89,705 | | | | 11.60% | |  | | |
| Race | | | | | | |  | | |  | | | <0.001 |
| Asian | | 2,060 | | 1.57% | 14,830 | | | | 1.92% | |  | | |
| Black | | 11,619 | | 8.87% | 61,663 | | | | 7.97% | |  | | |
| White | | 98,261 | | 74.97% | 562,973 | | | | 72.78% | |  | | |
| Other | | 8,262 | | 6.30% | 44,930 | | | | 5.81% | |  | | |
| Unknown | | 10,860 | | 8.29% | 89,153 | | | | 11.53% | |  | | |
| Ethnicity | | | | | | |  | | |  | | | <0.001 |
| Hispanic | | 5,384 | | 4.11% | 47,567 | | | | 6.15% | |  | | |
| Not Hispanic | | 95,902 | | 73.17% | 557,312 | | | | 72.05% | |  | | |
| Unknown | | 29,776 | | 22.72% | 168,670 | | | | 21.80% | |  | | |
| Geographic Region | | | | | | |  | | |  | | | <0.001 |
| West | | 5,666 | | 4.32% | 72,435 | | | | 9.36% | |  | | |
| Southeastern | | 42,134 | | 32.15% | 236,944 | | | | 30.63% | |  | | |
| Southcentral | | 26,208 | | 20.00% | 123,032 | | | | 15.90% | |  | | |
| Northeast | | 48,986 | | 37.38% | 166,773 | | | | 21.56% | |  | | |
| Northcentral | | 6,273 | | 4.79% | 129,938 | | | | 16.80% | |  | | |
| Unknown | | 1,795 | | 1.37% | 44,427 | | | | 5.74% | |  | | |
